# Estimating Treatment Effects for Time-to-Treatment Antibiotic Stewardship in Sepsis

**DOI:** 10.1101/2022.08.29.22279330

**Authors:** Ruoqi Liu, Katherine H. Buck, Jeffrey M. Caterino, Ping Zhang

## Abstract

Sepsis is a life-threatening condition with high in-hospital mortality rate. The timing of antibiotic (ATB) administration poses a critical problem for sepsis management. Existing work studying antibiotic timing either ignores the temporality of the observational data or the heterogeneity of the treatment effects. In this paper, we propose a novel method to estimate **T**reatmen**T** effects for **T**ime-to-**T**reatment antibiotic stewardship in sepsis (**T4**). **T4** estimates individual treatment effects (ITEs) by recurrently encoding temporal and static variables as potential confounders, and then decoding the outcomes under different treatment sequences. We propose a mini-batch balancing matching that mimics the randomized controlled trial process to adjust the confounding. The model achieves interpretability through a global-level attention mechanism and a variable-level importance examination. Meanwhile, we incorporate **T4** with uncertainty quantification to help prevent overconfident recommendations. We demonstrate that **T4** can identify effective treatment timing with estimated ITEs for antibiotic stewardship on two real-world datasets. Moreover, comprehensive experiments on a synthetic dataset exhibit the outstanding performance of **T4** compared to the state-of-the-art models on ITE estimation.

## Introduction

Sepsis is the body’s overwhelming response to infection, which can lead to tissue damage, organ failure, amputations, and death. Sepsis contributes to 6% of hospitalizations and 35% of in-hospital deaths^1^, and costs more than $27 billion annually in the USA^2^. Based on a recent study on Medicare beneficiaries, approximately 30% of septic patients do not survive for 6 months^3^. Broad-spectrum antibiotics are the first-line medications for sepsis^4,5^ because bacterial infection causes most cases^6^.

The current sepsis treatment guideline for antibiotic timing is a one-size-fits-all approach, and when patients with suspected sepsis should receive antibiotics remains controversial^7,8^. The Surviving Sepsis Campaign (SSC) recommends initiating broad-spectrum antibiotics within 1 hour for any patient with suspected sepsis or septic shock^9,10^. While the recommendation is supported by several large observational studies^11,12,13^, there is substantial concern that striving for 1-hour antibiotic delivery for all patients with suspected sepsis may cause serious harm (e.g., antibiotic resistance, C. difficile infection)^8,14,15^. Thus determining personalized antibiotic timing at the bedside is urgently needed.

Computational algorithms^16,12,13,17^ have been leveraged for examining the optimal antibiotic timing for septic patients using electronic health records (EHRs). EHRs contain irregularly sampled temporal data, including patient’s lab test results, vital signs and demographics. The main idea is to estimate the treatment effects of different timings of antibiotics on septic outcomes (e.g., in-hospital mortality). However, most studies have either ignored the temporality of EHRs^12,13^ or the heterogeneity of treatment effects^16,17^. These two issues are crucial for identifying effective and precise therapy for septic patients.

Some other literature^18,19,20,21,22^ of treatment design for sepsis treat the problem as an off-policy evaluation using observational data. For example, Komorowski et al.,^21^ propose a model to learn treatment policy based on patient trajectories (i.e., states, actions and observations) by optimizing a reward determined by patient survival. However, our method derives optimal treatment options by estimating individual potential outcomes for future timestamps. Our problem setting is more challenging that 1) we need to do counterfactual reasoning based on only observed data and 2) we need to adjust timevarying confounding and estimate unbiased individual causal effects.

In this paper, we study the problem of identifying the most effective timing for antibiotic administration in septic patients using EHR data. As shown in Figure 1, the patient’s information are extracted and compiled from EHRs and then used to build the model for antibiotic administration timing recommendation. To address the aforementioned challenges, we propose a novel framework to estimate **T**rustworthy **T**reatment effects for **T**ime-to-**T**reatment antibiotic stewardship in sepsis (**T4**). The proposed model **T4** first estimates individual treatment effects (ITEs) of receiving antibiotics by recurrently encoding temporal and static information obtained before the current timestamp (baseline period), and then decoding the potential outcomes under different treatment sequences after the current timestamp (follow-up period). We apply balancing matching for each mini-batch via treatment propensity as balancing scores to construct a *pseudo* balanced mini-batch, thus adjust the influence of confounders. We also provide the model interpretability of treatment recommendation by analyzing: 1) the contribution of each timestamp in the baseline to treatment recommendation with attention mechanism and 2)the contribution of each variable to treatment recommendation via variable importance examination that excludes each variable in evaluating the influence on model loss. Meanwhile, we adopt MC Dropout^23^ to estimate uncertainty and quantify the confidence behind the ITE estimation and treatment recommendation.

**Figure 1.**
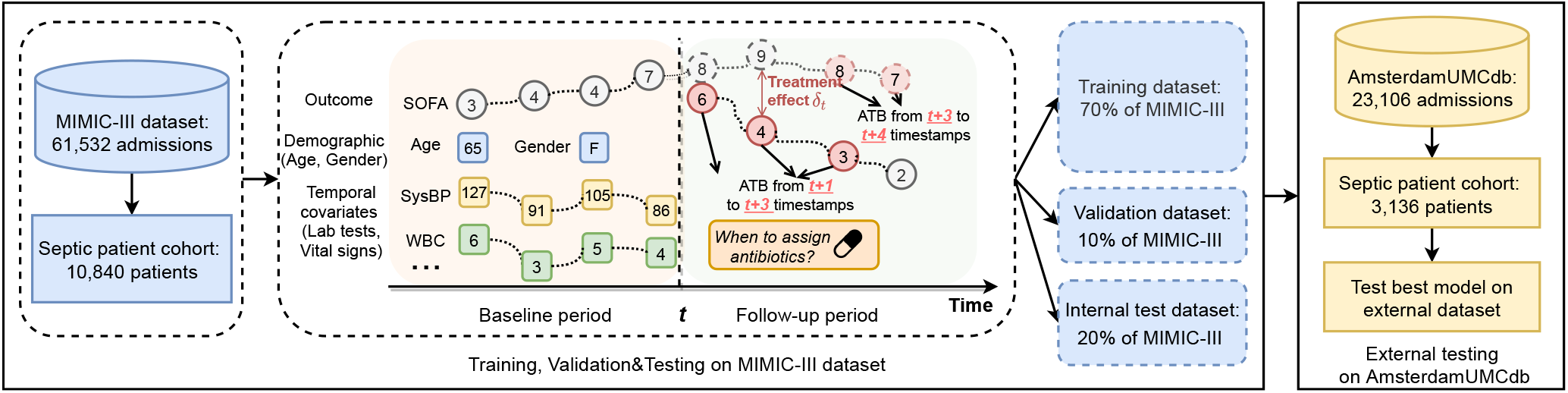
Overall data flow of **T4** framework. The data from MIMIC-III is randomly split into training, validation and testing datasets with percentages of 70%, 10% and 20%, respectively. The validation dataset is used to select the best model parameters and the testing dataset is used as an internal evaluation dataset. **T4** framework is used to estimate individual treatment effects for ATB administration timing recommendation. An external dataset obtained from AmsterdamUMCdb is used as an external test set.

We evaluate the effectiveness of treatment recommendation on two nonoverlapping real-world EHR datasets: Medical Information Mart for Intensive Care version III (MIMIC-III)^24^ and AmsterdamUMCdb^25^. The results show that the mortality rate of patients who receive the antibiotics at the time we recommend is notably lower than the patients who do not, indicating that our model offers effective timings of antibiotic administration that help to reduce the mortality rate. We demonstrate the application of our model on time-to-treatment recommendation using a concrete patient example. We also analyze model interpretability by visualizing the global and variable-level contribution to treatment recommendation via a concrete case study. Moreover, we conduct comparison experiments on ITE estimation using a synthetic dataset, and our model outperforms the state-of-the-art ITE estimation methods.

The contributions of this paper include the following:

- We propose an end-to-end treatment timing recommendation framework that seamlessly integrates the treatment effect estimation model, uncertainty quantification, and model interpretability for making transparent treatment recommendation.
- We develop a new ITE estimation method that can model time-varying information and adjust the influence of temporal confounding variables via balancing matching that mimics the randomized controlled trial process.
- We incorporate the ITE estimation with uncertainty quantification and interpretable analysis to achieve reliable treatment recommendation.
- We illustrate the usage of the proposed model in two real-world EHR datasets. The results show that our model can successfully identify effective timing of treatment and thus pave the way for personalized and precision medicine. We further conduct comprehensive comparison experiments on a synthetic dataset for ITE estimation.

## Overall framework

**T4** recurrently encodes the patient’s temporal covariates extracted from the baseline period, then decodes the potential outcomes with different treatment sequences in the follow-up period. Both encoder and decoder are built based on long short-term memory (LSTM)^26^, a deep recurrent neural network that is widely used for modeling time series data. **T4** adjusts the influence of confounders via *balancing matching* (Fig. 3) to generate balanced mini-batches. Each patient in the mini-batch is matched with the corresponding counterfactual outcomes using the observed outcomes of his or her similar patients in different treatment groups. The similarity of patients are estimated using propensity scores^27^ of receiving the current treatment. Figure 2 illustrates the framework of the proposed method. The training procedure of **T4** is shown in Algorithm 1.

**Figure 2.**
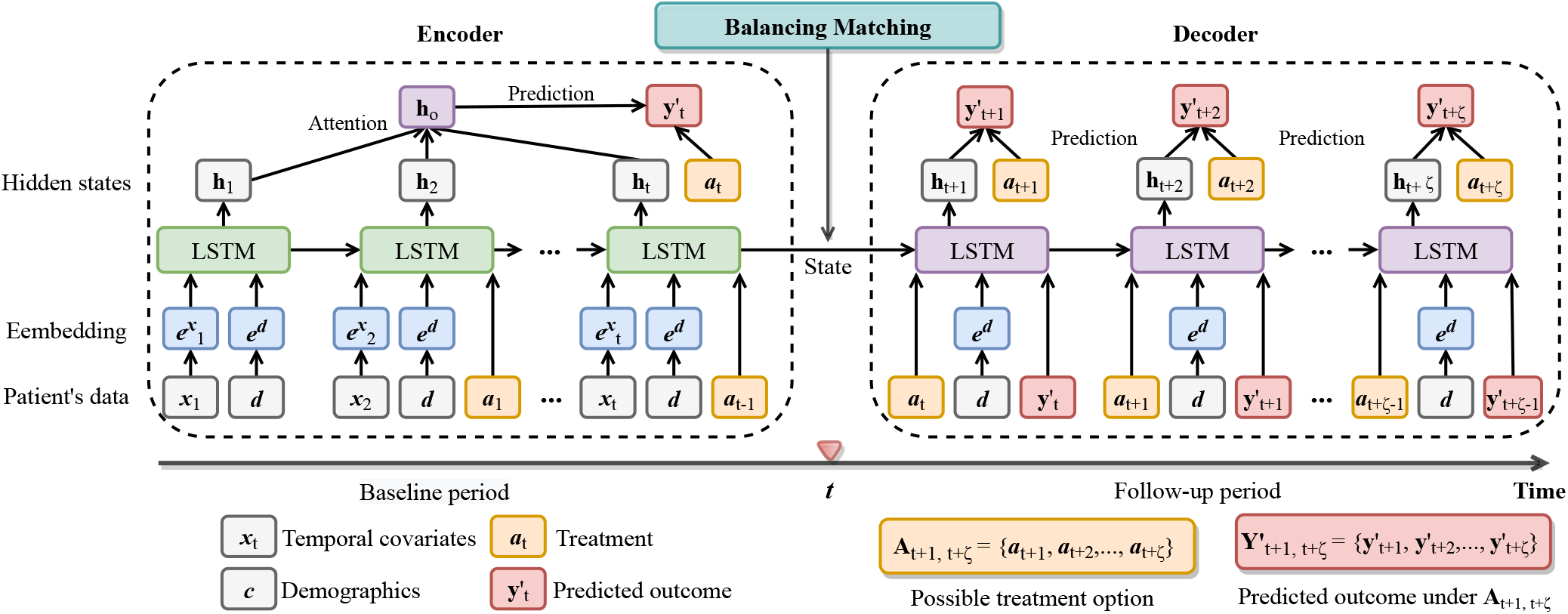
The framework of **T4. T4** consists of three main components: 1) the encoder network recurrently encodes the patient’s baseline information, including temporal and static covariates, and treatment assignments via the LSTM network; 2) the decoder network is initialized with encoder outputs and predicts the outcomes under different treatment sequences; 3) the balancing matching constructs balanced mini-batches via propensity as balancing scores during the training process. The details of balancing matching are shown in Fig. 3.

**Figure 3.**
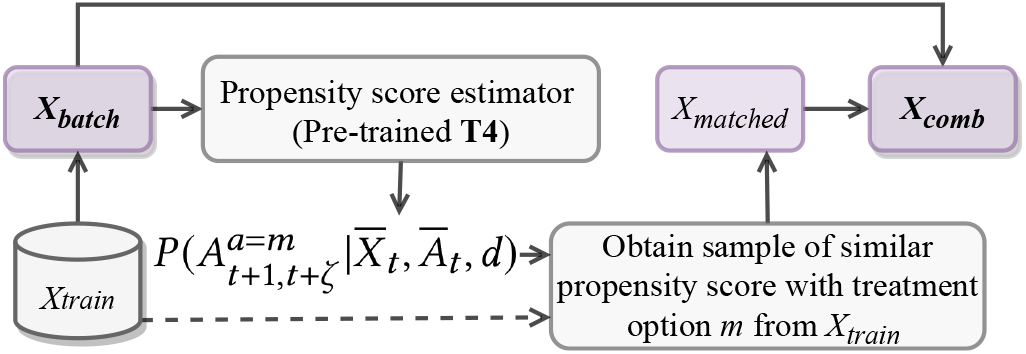
Illustration of the balancing matching. During the training process, each patient in the batch is matched with the corresponding counterfactual outcomes using the observed outcomes of his/her nearest neighbors in other treatment groups in the training data. A pre-trained **T4** is used to estimate the propensity scores for computing the patient’s distance. The matched batch and original batch are combined together for the training.

### Algorithm 1

**T4** Training Procedure

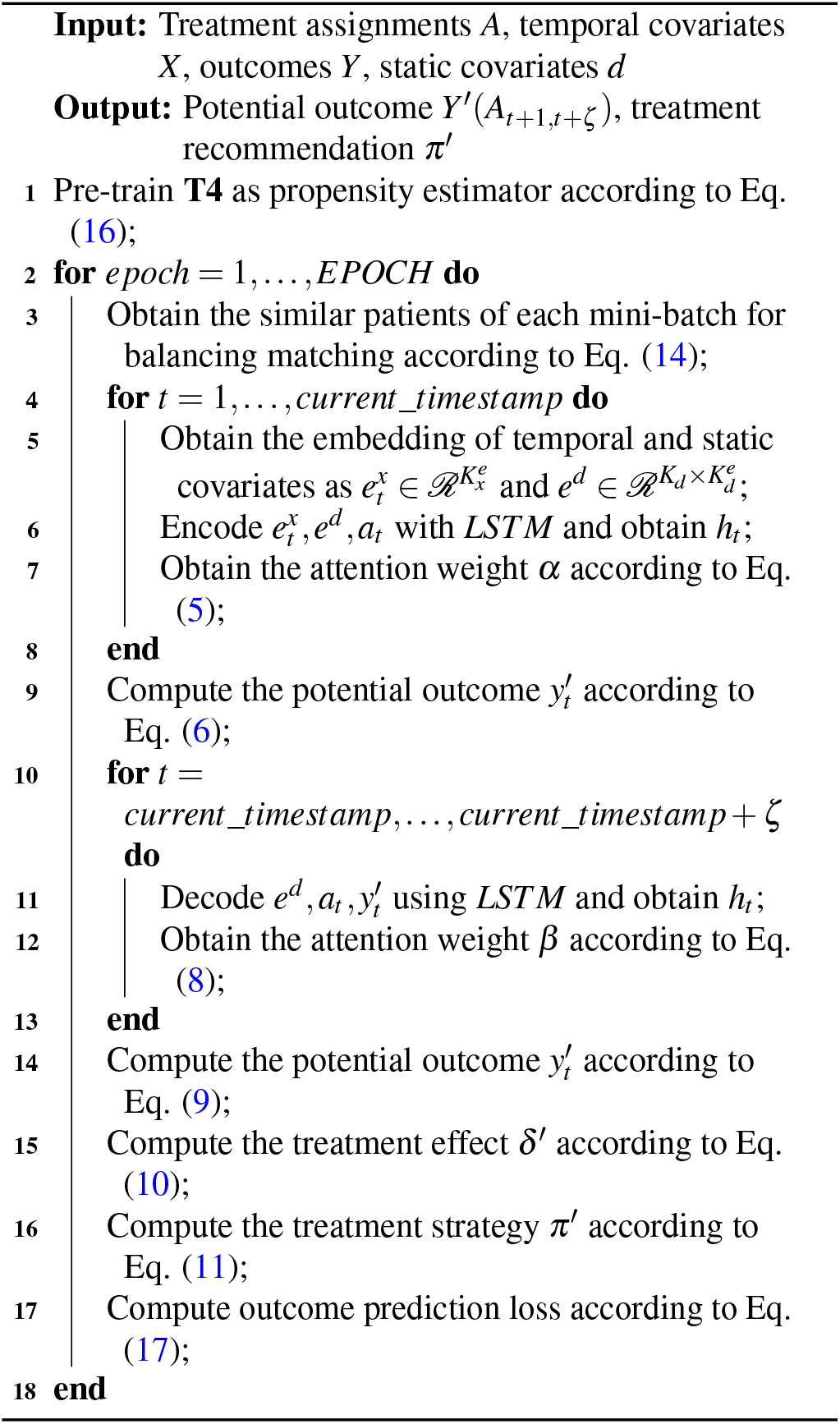

## Results

### Datasets

#### MIMIC-III

MIMIC-III^24^ is a large, freely-available database comprising de-identified health-related data associated with over 40,000 patients who stayed in critical care units of the Beth Israel Deaconess Medical Center between 2001 and 2012. It contains patients’ demographics, vital signs, lab tests and treatment assignments.

#### AmsterdamUMCdb

AmsterdamUMCdb^25^ is the first freely accessible European intensive care database. It is endorsed by the European Society of Intensive Care Medicine (ESICM) and its Data Science Section. It contains de-identified health data related to tens of thousands of intensive care unit admissions, including demographics, vital signs, laboratory tests and medications.

In both datasets, we included adult septic patients fulfilling the international consensus sepsis-3 criteria^28^. We extracted 10,840 patients and 3,136 patients from MIMIC-III and AmsterdamUMCdb after applying exclusion criteria respectively. Here, the causal inference problem we studied is the treatment effects of antibiotic therapy among sepsis patients given the observed confounding variables. There are three essential components that should be identified from the patient data: (1)**Treatments**: we consider multiple kinds of antibiotic therapy during the ICU stays. At each timestamp, a binary treatment indicator will denote whether the patient is assigned with antibiotics or not; (2) **Confounders**: we obtain 22 temporal covariates (i.e., vital signs: temperature, heart rate, etc.; lab tests: potassium, sodium, etc.) and 4 static covariates (i.e., age, gender, etc.) as potential confounders; (3) **Outcomes**: we compile the 24-hour Sepsis-related Organ Failure Assessment (SOFA)^29^ score as the primary outcome, which is computed based on the degree of dysfunction of six organ systems. The definition of sepsis-3 patient cohort is in study design of Methods, the list of antibiotics is in Supplementary Table 1, the list of patient’s covariates is in Supplementary Table 3 and computation of SOFA scores is in Supplementary Table 2.

### Model performance

#### Population level analysis

As we have no access to the counterfactual outcomes in the real-world dataset, so that we are not able to directly evaluate the model performance in terms of counterfactual prediction. Thus, we evaluate the model performance by comparing the treatment effects of recommended timing of administration (determined by estimated ITE according to Eq. (11)) and the observed timing of administration on patients’ mortality rate. Specifically, we first obtain a target group of patients whose observed timing of ATB administration is different from the model recommendation. Then we derive a compared group to the target group by involving the most similar patients whose observed timing of ATB administration matches the model recommendation. We use the variables obtained from the baseline period (time window before the follow-up period) to perform patient similarity. We use Euclidean distance as the similarity measurement. Finally we compare the mortality rate within these two groups and expect that the mortality rate of patients whose observed treatments match our recommendation would be much lower than the patients whose observed treatments are different from recommendation.

As shown in Fig. 4, we compute and compare 30 days mortality rate and 60 days mortality rate on two datasets, respectively. The black dashed lines in the two plots denote the average mortality rate among the population, which are the baselines for the comparison. We find that the mortality rate of patients who receive treatments at different timestamps as our recommendation is higher than the average mortality rate baseline, while the mortality rate of patients who receive the treatments at the same timestamps as our recommendation is lower than the baseline. We evaluate the model concerning different lengths of the follow-up period (i.e., *ζ* ∈ { 3, 4, 5, 6, 7) } and the mortality rates for patients with the same treatments are consistently lower than the mortality rates for patients with different treatments. Results show that our model recommends effective treatment strategies (reflecting on lower mortality rate), and provides potential clinical insights for doctors to decide the timing of antibiotic administration for the septic patients. We also observe that model performs consistently on the external testing set from AmsterdamUMCdb, which demonstrates the robustness of our model when applying to a different dataset with different feature distribution.

**Figure 4.**
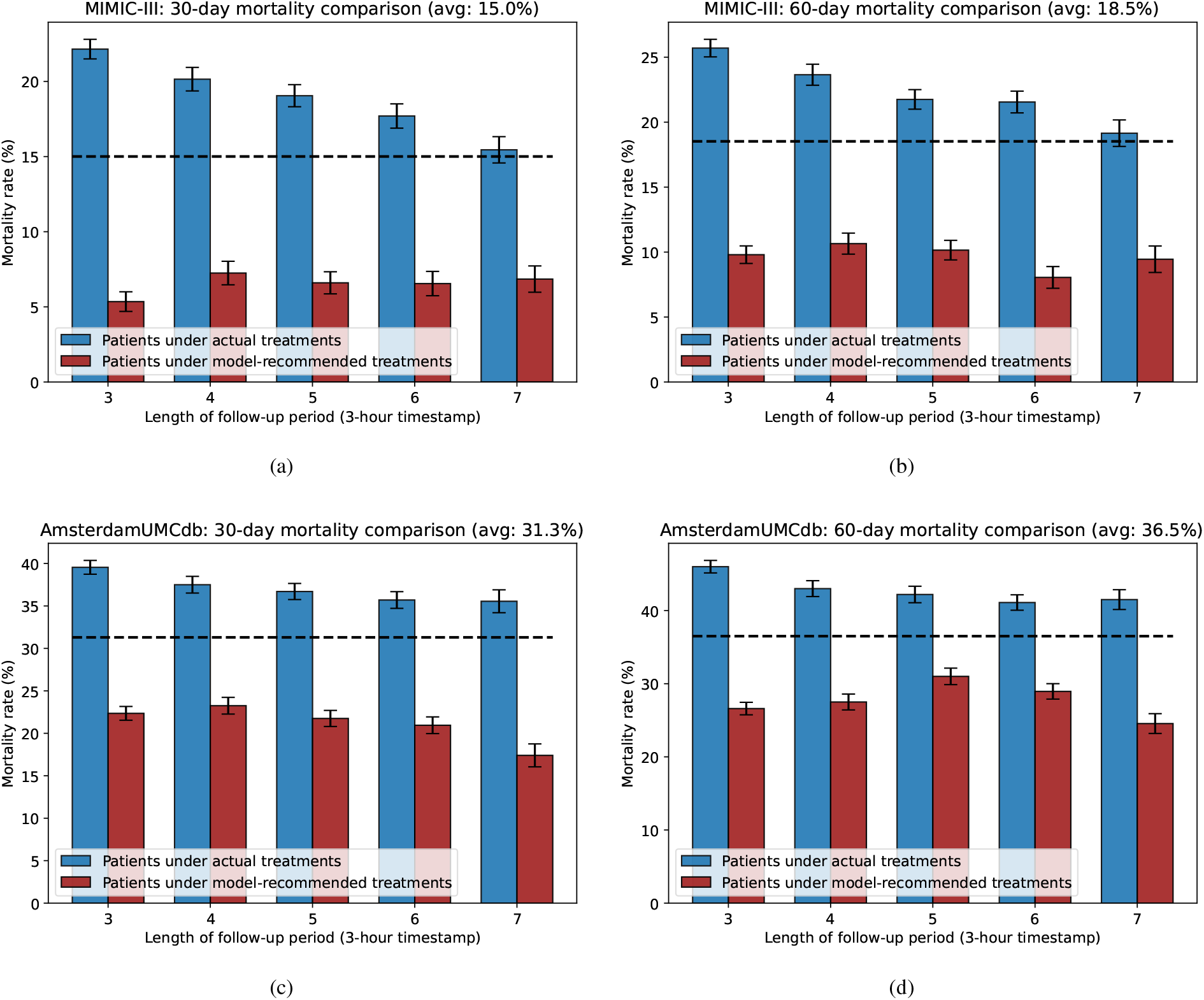
Mortality rate comparison of two datasets. Within the bar-chart, the error bars denote 95% confidence interval with *n*=30 bootstrap samples. Blue and red bars denote patients under actual treatments and patients under model-recommended treatments, respectively. The total mortality rate of two groups of patients is plotted using the black dashed line, which serves as the baseline. Fig. 4(a) and 4(b) are mortality comparison on MIMIC-III dataset. Fig. 4(c) and Fig. 4(d) are mortality comparison on AmsterdamUMCdb dataset.

#### Individual level analysis

To further demonstrate how our model recommends antibiotics based on the estimated ITEs with uncertainty quantification, we utilize a real-world patient case. As shown in Fig. 5(a), we use the predicted ITEs (red line) equipped with uncertainty estimates (red shadowed area) as the criteria to recommend the timing of antibiotic administration at each timestamp. Specifically, at each timestamp, we recommend the antibiotics if the upper bound of the predicted ITE is lower than zero, and we will not recommend if the lower bound of the predicted ITE is higher than zero. Here, zero serves as the baseline, denoting no difference between having a treatment and not. In this example, the upper bound of the predicted ITE is lower than zero for all the timestamps in the follow-up period. Thus, the optimal antibiotic recommendation provided by our model is to continuously take antibiotics during the follow-up period.

**Figure 5.**
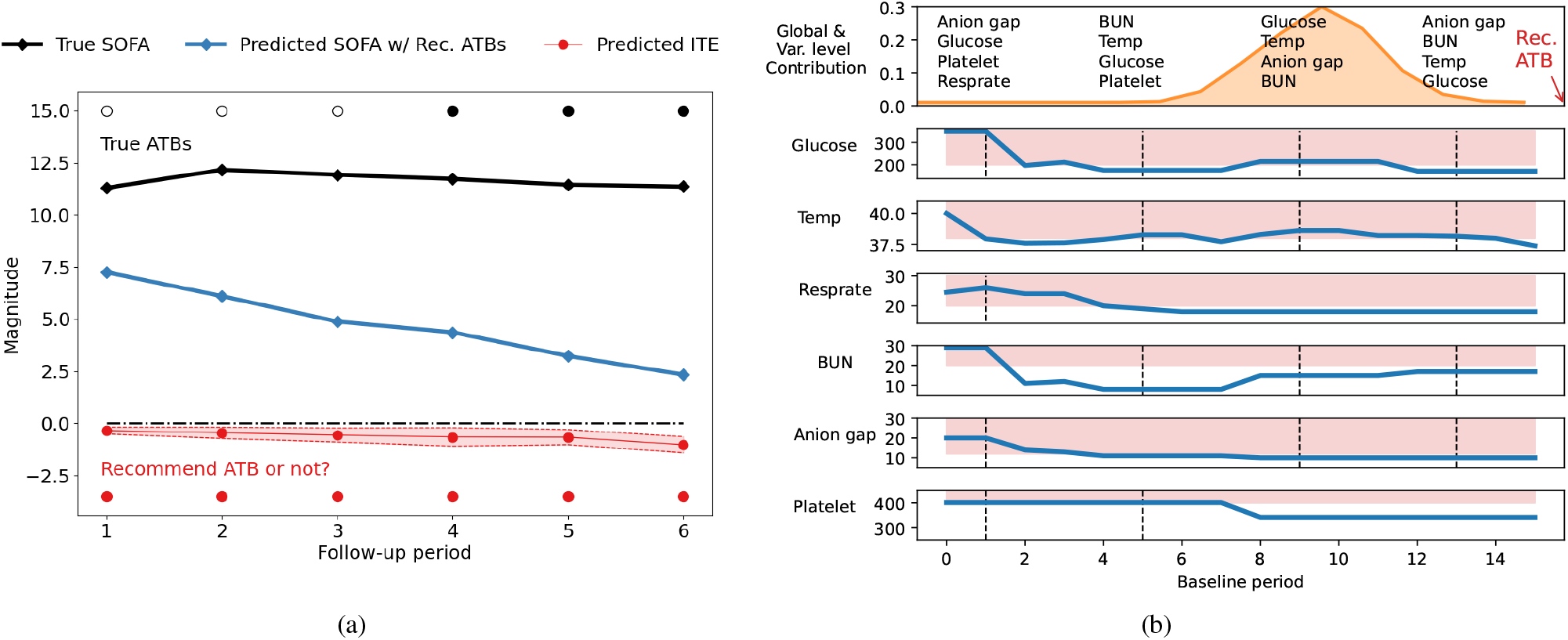
The case study of a patient for treatment recommendation and model interpretability. In Fig. 5(a), the predicted values of ITEs (red line with shadowed area denoting the uncertainty estimates) for antibiotic (ATB) recommendation. An ATB will be recommended to the patient if the upper bound of predicted ITE is lower than zero and will not be recommended if the lower bound of predicted ITE is higher than zero, where zero is the threshold for determining whether to recommend ATBs. In Fig. 5(b), the most important global baseline timestamps (orange area) and variables contributing to the treatment recommendation are denoted at the top subplot. An ATB is recommended at the end of the baseline period.

We also compare true outcomes under the actually received antibiotics and predicted outcomes under the recommended antibiotics. Here, we use the SOFA score as the outcomes, where higher values are associated with worse status and higher mortality rate^30,31^. From Fig. 5(a), we observe that the patient receives antibiotics since the 4-*th* timestamp, which results in large SOFA scores during the follow-up period. Conversely, our model recommends the patient to take antibiotics earlier (from the 1-*th* to the 6-*th* timestamp), and the predicted SOFA scores under recommended timing of antibiotics are much lower than the true SOFA scores. The results demonstrate that our model can identify effective timing of antibiotic administration for septic patients to help improve their disease condition and reduce the mortality rate.

#### A Case Study for Model Interpretability

We demonstrate the model interpretability of treatment recommendation using a concrete case study. We visualize both global and variable level contribution in Fig. 5(b). We also plot in this figure the dynamics of each important variable. Here, the vital signs include temperature, and respiratory rate (resprate); the lab tests include glucose, blood urea nitrogen (BUN), anion gap, and platelet. We use the dashed black lines to denote the timestamps with high contribution to the treatment recommendation. We observe that the values of most variables are within or close to the abnormal range at those high contribution timestamps. Our model recommends the patient to take antibiotics at the end of the baseline period with regards to these warning signals. Taking the glucose as an example, the normal range should be lower than 140 mg/dL, and a reading of more than 200 mg/dL indicates diabetes^1^. We observe that the values of glucose are far from the nor-mal range, which maintain above 200 mg/dL and even reach 300 mg/dL at early stage. This case study shows that our model achieves the transparent treatment recommendation via visualizing important timestamps and variables contributing to the recommendation, paving the way for interpretable and precise treatment recommendation.

## Discussion

In this study we propose a novel framework to estimate treatment effects for treatment recommendation. The proposed model **T4** first estimates ITEs by recurrently encoding historical temporal patient information and static information, and then decoding the potential outcomes under different treatment sequences. We apply balancing matching for each minibatch to construct a balanced mini-batch and adjust the influ-ence of confounders. We also provide the model interpretability of treatment recommendation to analyze both global-level and variable-level contribution via attention mechanism and variable importance analysis, respectively. Meanwhile, the model uncertainty quantification helps to avoid overconfident treatment recommendation. We illustrate the usage of the proposed model in two real-world EHR datasets, showing that our model can successfully identify effective treatment strategies and thus pave the way for personalized and precision medicine.

### Individual treatment effect estimation

#### Comparison experiments

Several studies have proposed estimating individual treatment effects (ITEs) using causal inference techniques on observational data, such as matching-based methods (e.g., propensity score matching^27^), forest-based methods (e.g., causal forest^32^), or representation learning-based methods (e.g., counterfactual regression^33^). These methods are mainly designed for static data and are not satisfied for estimating ITEs on EHRs, while the proposed **T4** fully considers the time-varying information and adjust the temporal confounding via balancing matching operation.

To illustrate the model performance on ITE estimation and treatment recommendation, we design experiments on a synthetic dataset. We simulate 5000 patients with 50 timestamps, 20 temporal covariates and 5 static covariates. We use the first 40 timestamps as baseline period and the remaining as follow-up period. We use Precision in Estimation of Heterogeneous Effect (PEHE) and the error of Average Treatment Effect (*ε*ATE) to evaluate the model performance. We conduct comparison experiments against the state-of-the-art methods of ITE estimation: (1) **Classical methods:** Linear Regression (LR)^34^, Random Forest (RF)^35^ and support vector machine (SVM)^36^; (2) **Forest-based methods:** Causal Forest (CF)^32^ and Bayesian Additive Regression Trees (BART)^37^; (3) **Representation learning-based methods:** Counterfactual Regression (CFR)^33^, GANITE^38^ and Dragonnet^39^. (4) **Time-varying based methods:** Recurrent Marginal structural Network (RMSM)^40^ and Counterfactual Recurrent Network (CRN)^41^.

Results in Supplementary Table 8 show that our model outperforms the state-of-the-art ITE estimation methods.

#### Ablation study on balancing matching

We evaluate the influence of different percentages of balancing matching samples on the model performance. We vary the percentages from 0 to 100% and show the performance change in Supplementary Fig. 8. We observe that the error of ITE estimation in terms of both PEHE and *ε*ATE decreases as including more matching samples in a mini-batch during the training process. Specifically, performance highly increases when only a small number of matching samples (around 20%) are provided, and the curve tends to gently slope downward as the percentage of matching samples exceeds 30%. The results demonstrate that the balancing matching improves the model performance on ITE estimation by largely decreasing the estimation error.

### Controversies in antibiotics for septic patients

The timing of antibiotic treatment is controversial. While many studies suggest early antibiotic regimens for any patients with suspected sepsis or septic shock, there is substantial concern that early antibiotic assignments may cause serious harm including higher mortality rates^7^. Several recent cohort studies and RCTs^8,15,42,43,44^ suggest that the timing of antibiotic treatment should depend on the severity of illness (i.e, sepsis, severe sepsis or septic shock) and the likelihood of true infection. They point out that immediate antibiotic regimens benefit patients with severe illness (e.g., septic shock) while in less critically ill patients, immediate antibiotic regimens may lead to overprescribing and potential harm. For example, a recent study^45^ shows that an overdose of antibiotics is associated with a 20% increase in the odds of death in patients who received adequate therapy. According to the study^45^, the morbidity of overdose antibiotics may be more obvious in less critically ill patients compared to the patients with septic shock as the morbidity of other acute severe illness surpasses the possible morbidity comes from an antibiotic overdose. There are also some general explanations for the association between antibiotic overdose and higher mortality. Besides the antibiotic resistance, antibiotics themselves also cause harm (e.g., organ injury, mitochondrial dysfunction, the impact on the microbiome, and overgrowth by fungi and Clostridium difficile)^46^. Specifically, the study^45^ shows that unnecessarily broad empiric therapy was associated with a 26% increased risk of Clostridium difficile infection. Thus determining personalized antibiotic timing at the bedside is urgently needed.

### Limitation of public clinical data

#### Unavailability of counterfactual outcomes

As the ground truth counterfactual outcomes are not available in the real-world data, we evaluate the effectiveness of our model in two ways: 1) mortality rate comparison as shown in Fig. 4 and 2) factual prediction on SOFA scores as shown in Supplementary Table 7.

#### Type of antibiotic treatment

The choice of the type of antibiotic treatment is based on suspected infection sites according to empirical antibiotic studies and guidelines^47,48^. However, the sites of suspected infection are not available in our public clinical datasets (MIMIC-III and AmsterdamUMCdb), especially during the first 48 hours since ICU admission as the determination of true infection sites is complicated and takes time to obtain the results. In the future, we will incorporate suspected infection sites to provide recommendations for a specific type of antibiotic, orthogonal etc.

#### Blood cultures

Blood cultures are deemed as the gold standard for antibiotic treatment regimens (e.g., initiation or de-escalation of antibiotics). However, in our public datasets, most blood cultures are taken 12 hours within the ICU admission and usually the results are available after 2-3 days. According to the proposed framework as illustrated in 2, we only leverage the first 48 hours data since ICU admission and therefore the results of blood cultures may not be available in this period.

In future work, we can develop a more practical and precise antibiotic recommendation system that combines the model’s general recommendations with real patient conditions (e.g., suspected infection sites, blood cultures and other concomitant therapies) if available.

In summary, we propose a computational framework to estimate treatment effects for time-to-treatment antibiotic stewardship in sepsis. Experiments on both real-world and synthetic datasets have demonstrated the superiority of the model performance compared to the state-of-the-art models, while also providing interpretability of the treatment recommendation.

## Methods

In this section, we first introduce the study design, then we present the proposed model for estimating treatment effects.

### Study design

We evaluate the proposed treatment recommendation framework through a retrospective study on two large real-wrold EHR datasets (MIMIC-III and AmsterdamUMCdb) with recorded patients’ demographics, vital signs, lab tests, medications and diagnosis.

#### Definition of Sepsis in two datasets

We obtain the septic patients according to the recent sepsis-3 criteria^28^ with respect to **1)** *t*_*susp*_: time of clinical suspicion of infection (i.e., earlier timestamp of antibiotics and blood cultures within a specified duration) and **2)** *t*_*SOFA*_: two-point deterioration in SOFA score^29^ within a 6-hour period. The patient is diagnosed with sepsis when these two events happen close to each other. Specifically, *t*_*SOFA*_ happens 24 hours earlier than *t*_*susp*_ or 12 hours later than *t*_*susp*_. We exclude patients whose age is under 18 years old at the time of ICU admission. We also exclude patients whose ICU stay is less than 9 hours or longer than 20 days.

We extract 22 temporal covariates (i.e., vital signs and lab tests) and 4 static covariates (Supplementary Table 3). We encode the patients’ time series data into discrete 3-hour time steps. The covariates with multiple records within a single time step are averaged. The missing data are imputed using the values obtained from the last time step.

### Preliminary

We extract patient information from longitudinal observational data. For each patient, let *Ā*_*T*_ = {*a*_1_, *a*_2_, …, *a*_*T*_} ∈ *𝒩* ^*T*^ be the treatment assignments with *a*_*t*_ = 1 if the patient receive the treatment at *t*-th timestamp and *a*_*t*_ = 0 otherwise. Let 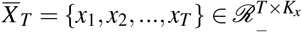 be the temporal covariates and 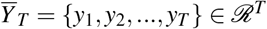 be the outcomes of *T* timestamps. The patient has static covariates 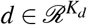, such as gender and age. The observational data for the patient can be represented as 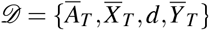.

Our goal is to estimate the treatment effects with temporal and static covariates via predicting the potential outcomes under different treatment sequences. We adopt the potential outcomes framework^49,50^ to examine the causal effects under the treatments. The potential outcome is the outcome that would have been observed if the patient had received treatment. We extend the potential outcome framework in our application scenario. Given the observational data up to *t*-th timestamp and treatment assignments **A**_*t*+1,*t*+*ζ*_, the patient has potential outcomes 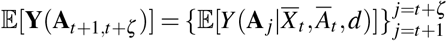 in the following *ζ* time period. Specifically, **A**_*t*+1,*t*+*ζ*_ = {*a*_*t*+1_, *a*_*t*+2_, …, *a*_*t*+*ζ*_} denotes any treatment assignments from *t* + 1 to *t* + *ζ* timestamp. For each timestamp *j*, there are two potential outcomes 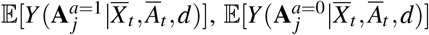, which are corresponding to different treatment assignments.

To estimate the treatment effect of a given treatment assignment during the following *ζ* time period, we define the individual treatment effect (ITE), *δ* _*j*_ on (*t* + *j*)-th timestamp as follows,

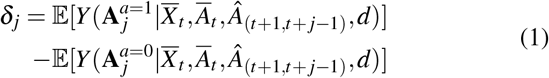

where 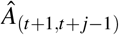 is the learned optimal treatments between timestamp *t* + 1 and *t* + *j* − 1. The treatment effects of *ζ* time period is Δ = [*δ*_*t*+1_, …, *δ*_*t*+*ζ*_]. In this paper, we use the Sepsis-related Organ Failure Assessment (SOFA) scores^29^ (i.e., range from 0 to 24, larger values associated with severe disease status and higher mortality) as outcomes. The computation of SOFA scores can be found in Supplementary Table 2. The recommended treatment assignments are as follows,

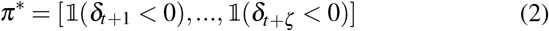

where 𝟙(·) equals to 1 if the inside expression is true otherwise 0.

### Assumptions

Our ITE estimation is based on the standard causal assumptions^51,52^ as follow,

#### Assumption 1

**(Consistency)** *The potential outcome under treatment history Ā*_*t*_ *equals to the observed outcome if the actual treatments history is Ā*_*t*_.

#### Assumption 2

**(Positivity)** *Given the observational data of the history, if the the probability* 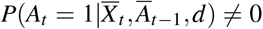, *then the probability of receiving treatment* 0 *or* 1 *is positive, i*.*e*.,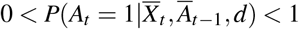, *for all Ā*_*t*_.

#### Assumption 3

**(Sequential Strong Ignorability)** *Given the observational data of the history, the treatment assigned for time t is independent of the potential outcome of time t, i*.*e*., 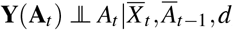, *for all treatment sequences* **A**_*t*_.

### Treatment effect estimation with T4

**T4** recurrently encodes the patient’s temporal covariates via Long short-term memory (LSTM)^26^, then decodes the potential outcomes with different treatment sequences. **T4** adjusts the influence of confounders via balancing matching to generate balanced mini-batches. Figure 2 illustrates the framework of the proposed method.

#### Encoder for baseline period

We convert the initial high-dimensional covariates 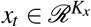 into a lower dimensional and continuous data embedding 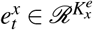 as,

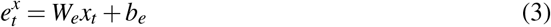

where 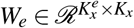 is the weight matrix, 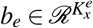 is the bias vector and 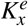 is the dimension of the embedded temporal vectors. That is, we have embedding of temporal covariates 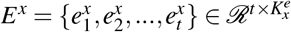. Similarly, we convert the static covariates (demographics) into embedding as 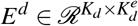, where *K*_*d*_ is the number of static covariates and 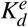 is the dimension of embedded static vectors.

Given the embedding of temporal and static covariates, and the treatment assignments at each timestamp, the encoder builds upon the LSTM as follows,

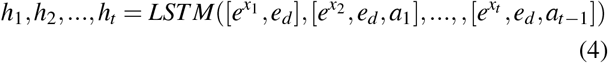

where 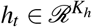 is the hidden state at *t*-th timestamp and *K*_*h*_ is the dimension of hidden vectors. The last hidden state *h*_*t*_ is used to initialize the decoder. We aggregated all the hidden states via attention mechanism for automatically focusing on important historical timestamps. We calculate the attention weight *α*_*t,s*_ using a method that concatenates each previous hidden state *h*_*s*_ with the current state *h*_*t*_, and the product of two states. That is,

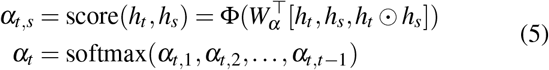

where Φ is hyperbolic tangent function, 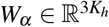 is learnable parameter matrix. Using the generated attention energies, we calculate the context vector *h*_*o*_ for each patient up to *t* time stamp as 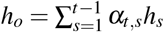.

We predict the potential outcome *y*_*t*_ using the attentively aggregated vector *h*_*o*_, current hidden state *h*_*t*_ and the treatment *a*_*t*_. The prediction serves as the input to the initial state of the decoder,

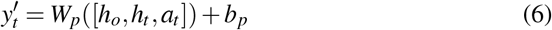

where 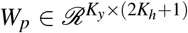 and 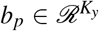 are parameters to learn.

#### Decoder for follow-up period

Initializing with the last hidden state of the encoder and true/predicted outcomes, the decoder recurrently predicts the potential outcome at each timestamp with different treatment sequences. We obtain the hidden states of decoder as,

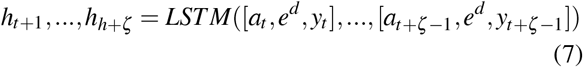

We integrate the encoder outputs and current hidden state of decoder via an attention layer. We generate the aggregated context vector *c*_*t*+ *j*_ at each timestamp as,

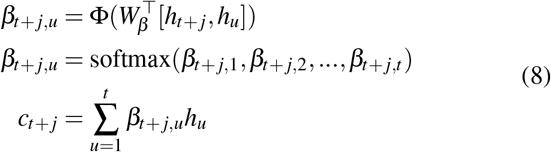

where Φ is hyperbolic tangent function, 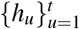 are the encoder outputs and 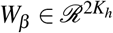 is the parameter to learn.

We predict the outcomes 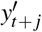 by combining the learned context vector with current treatment as

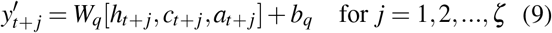

where 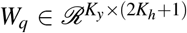 and 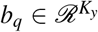 are parameters to learn. During the training, we use teach forcing technique with ratio equals to 0.5 to train the model with ground truth treatments and outcomes. In the inference/testing, we feed the decoder’s predictions (both outcomes and treatments) back to itself for each step. The current predictions are based on the previous predictions during the inference, which is consistent to the practical application scenario.

We identify the optimal treatment sequence via a greedy-style strategy instead of checking every possible treatment trajectory. We select the best treatment option according to the predicted outcomes at each step and use it for next prediction. Compared to permutation of all possible combination (up to 2^*ζ*^) of treatments, our strategy is more time efficient with the increasing of *ζ*.

Then we can compute the treatment effect for (*t* + *j*)-th timestamp using Eq.(1) as,

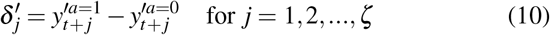

where 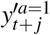 is the predicted outcome when receiving the treatment at (*t* + *j*)-th timestamp, and 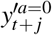 is the predicted outcome when not receiving the treatment. Thus we determine the optimal treatment assignments among all *ζ* time period using Eq.(2) as,

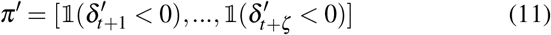

#### Balancing matching

To adjust the inherent treatment selection bias in the data, we adopt the idea of balancing scores^53^ to construct *pseudo* minibatches that mimic the corresponding randomized controlled trial (RCT) process (i.e., the treatment groups are randomly split and the patient distribution in each group is balanced.) We illustrate the process of balancing matching in Fig. 3. Specifically, we match, for each patient in the original minibatch, the unobserved counterfactual outcomes (i.e., the potential outcomes under other possible treatment options except the observed one), with the observed outcomes of nearest neighbors in the training data. There are several methods to obtain the nearest neighbor by computing the distance among individuals. Here, we estimate the distance via the propensity score^27^, which is defined as the conditional probability receiving the treatments *m*_*_ given historical information up to the current timestamp:

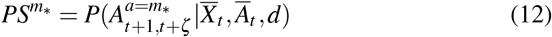

Here, *m*_*_ denote a possible treatment sequence during *ζ*. We use a pre-trained **T4** as a propensity score estimator to calculate the propensity scores for each patient in the training set.

The distance between patient *i* with treatments *m*_*_, and the patient *j* with treatments *n*_*_ is defined using absolute distance as,

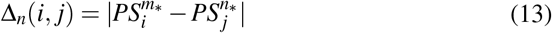

where 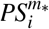 and 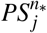 denote the estimated propensity scores for two respective patients. We then obtain the nearest neighbours of patient *i* in treatment group *n*_*_ as,

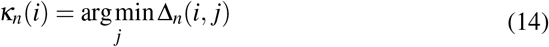

Finally the matched mini-batch is combined with the original mini-batch as a whole for the following training process.

#### Objective function

We first pre-train our model **T4** to estimate the propensity scores for balancing matching. We obtain the treatment prediction using a linear layer and sigmoid function as,

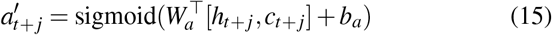

where 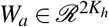 and *b*_*a*_ ∈ ℛ are parameters to learn. We use cross-entropy loss for the treatment prediction as,

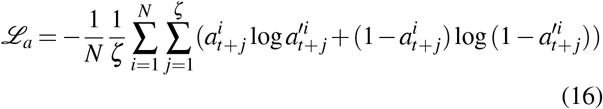

The training objective function for the outcome prediction is the mean squared error between the predicted potential outcomes and factual outcomes as,

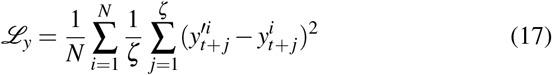

The overall training procedure of **T4** is demonstrated in 1.

### Model interpretability

Interpretability is a very desiring property in treatment effect estimation and treatment timing recommendation problems. In this paper, we realize the interpretability of treatment recommendation by analyzing both global-level contribution and variable-level contribution.

#### Global-level contribution

The global-level contribution is the contribution of each timestamp in the baseline period to the treatment recommendation given in the follow-up period. The outputs of the encoder are sent to the decoder and integrated together with the hidden states of decoder through the attention layer. We obtain the learned attention weights *β*_*t*+ *j,u*_ as the contribution of *u*-th timestamp to the treatment recommendation given at (*t* + *j*)-th timestamp according to Eq. 9.

#### Variable-level contribution

Each timestamp contains a number of temporal variables (e.g., lab tests, vital signs, etc.), and based solely on the contribution at the global level, we are unable to identify the impact of each individual variable. We then examine the contribution of each variable via a variable importance analysis. Specifically, given the temporal covariates *x*_*u*_, we first predict outcomes 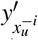 when excluding all the information from the *i*-th dimension of *x*_*t*_. Here, we mask the corresponding information by replacing them with the mean value of *i*-th variable across in the dataset. We compute the prediction loss 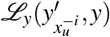 using Eq. (17) except that the predicted outcomes are replaced with 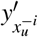. Finally, the contribution of each variable *i* at *u*-th timestamp is computed as,

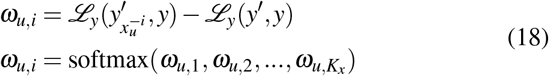

where ℒ_*y*_(*y*′, *y*) is the prediction loss when all features of *x*_*t*_ are included in the loss computation. We multiply the global-level contribution and the variable-level contribution (*β*_*t*+ *j,u*_*ω*_*u,i*_) to obtain the contribution of each variable at each timestamp.

### Uncertainty quantification

The uncertainty quantification of the estimated treatment effects is also important for treatment recommendation. In this paper, we adopt MC Dropout^23,54^ to quantify the model uncertainty by applying dropout during both training and testing process. Specifically, with the dropout enabled during the testing, the model generates a different output every forward pass for the same input. Suppose we have *K* iterations, and for iteration *k*, we obtain the estimated effect 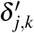. Then the model uncertainty 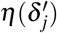 is computed as,

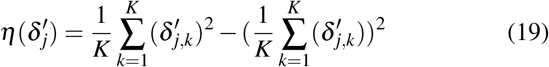

In this way, each ITE 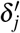 is equipped with according uncertainty estimates 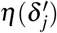. We use the estimated uncertainty for 1) quantifying the confidence associated with the estimated ITEs and provided recommendation. If the estimated uncertainty exceeds a certain threshold (i.e., 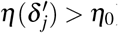), our model will alert the doctors that the provided recommendation are not reliable; 2) determining whether to assign a treatment at each timestamp. We derive the standard deviation from the variance and then calculate the 95% confidence intervals of ITE estimator. Our treatment recommendation strategy is that, at each timestamp in follow-up period, the treatment will be assigned to the patient if the upper bound of 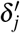 is less than zero, and the treatment will not be assigned if the lower bound of 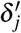 is larger than or equal to zero. The estimated uncertainty here is to enhance the robustness of prediction and guarantee the effectiveness of treatment recommendation.

### Implementation Details

The proposed model is implemented using Python 3.6 and PyTorch 1.4^2^ and trained on Ubuntu 20.04 with NVIDIA GeForce RTX 2080 Ti. We train our model using the adaptive moment estimation (Adam) algorithm. Dropout^23^ is enable during the training and testing for uncertainty estimation. The data is randomly split into training, validation and test sets with percentages of 70%, 10%, 20%, and the validation set is used to improve the models and select the best model hyper-parameters (Supplementary Table 4). We report the performance on the test sets for all methods. The final results are averaged on five random realizations. The values of both temporal and static covariates are normalized as follows,

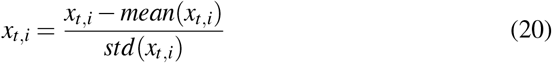

where *mean*(*x*_*t,i*_) and *std*(*x*_*t,i*_) are the mean and standard deviation of *i*-th variable in *x*_*t*_ over the entire dataset.

## Data Availability

MIMIC-III dataset is publicly available from PhysioNet (https://mimic.physionet.org/). AmsterdamUMCdb is publicly available from the Amsterdam Medical Data Science website (https://amsterdammedicaldatascience.nl/).

https://amsterdammedicaldatascience.nl/

## Data availability

MIMIC-III dataset is publicly available from PhysioNet^3^. AmsterdamUMCdb is publicly available from Amsterdam Medical Data Science website^4^.

## Code availability

The source code for this paper can be downloaded from the Github repository at https://github.com/ruoqi-liu/ T4 or the Zenodo repository at https://doi.org/10.5281/zenodo.7003982.

## Acknowledgements

This work was funded in part by the National Center for Advancing Translational Research of the National Institutes of Health under award number CTSA Grant UL1TR002733. The content is solely the responsibility of the authors and does not necessarily represent the official views of the National Institutes of Health.

## Author contributions

PZ conceived the project. RL and PZ developed the method. RL conducted the experiments. RL and PZ analyzed the results. RL, KB, JC and PZ wrote the manuscript. All authors read and approved the final manuscript.

## Additional information

Correspondence and requests for materials should be addressed to PZ.

## Footnotes

1 https://www.mayoclinic.org/diseases-conditions/diabetes/diagnosis-treatment/drc-20371451

1 https://pytorch.org/

1 https://mimic.physionet.org/

1 https://amsterdammedicaldatascience.nl/

## References

1. Rhee, C. et al. Incidence and trends of sepsis in us hospitals using clinical vs claims data, 2009-2014. Jama 318, 1241–1249 (2017).

2. Arefian, H. et al. Hospital-related cost of sepsis: a systematic review. J. Infect. 74, 107–117 (2017).

3. Buchman, T. G. et al. Sepsis among medicare beneficiaries: 1. the burdens of sepsis, 2012–2018. Critical care medicine 48, 276 (2020).

4. Treatment for sepsis. https://www.sepsis.org/sepsis-basics/treatment/ (2021).

5. Dellinger, R. P. et al. Surviving sepsis campaign: international guidelines for management of severe sepsis and septic shock, 2012. Intensive care medicine 39, 165–228 (2013).

6. Bacterial-infections in sepsis. https://www.sepsis.org/sepsisand/bacterial-infections/ (2021).

7. Moss, S. R. & Prescott, H. C. Current controversies in sepsis management. In Seminars in respiratory and critical care medicine, vol. 40, 594–603 (Thieme Medical Publishers, 2019).

8. Klompas, M. & Rhee, C. Current sepsis mandates are overly prescriptive, and some aspects may be harmful. Critical care medicine 48, 890–893 (2020).

9. Rhodes, A. et al. Campaña para sobrevivir a la sepsis: recomendaciones internacionales para el tratamiento de la sepsis y el choque septicémico: 2016. Crit Care Med [internet]. mar 45 (2017).

10. Levy, M. M., Evans, L. E. & Rhodes, A. The surviving sepsis campaign bundle: 2018 update. Intensive care medicine 44, 925–928 (2018).

11. Kalil, A. C., Johnson, D. W., Lisco, S. J. & Sun, J. Early goal-directed therapy for sepsis: a novel solution for discordant survival outcomes in clinical trials. Critical care medicine 45, 607–614 (2017).

12. Liu, V. X. et al. The timing of early antibiotics and hospital mortality in sepsis. Am. journal respiratory critical care medicine 196, 856–863 (2017).

13. Seymour, C. W. et al. Time to treatment and mortality during mandated emergency care for sepsis. New Engl. J. Medicine 376, 2235–2244 (2017).

14. Force, I. S. T. Infectious diseases society of america (idsa) position statement: why idsa did not endorse the surviving sepsis campaign guidelines. Clin. infectious diseases: an official publication Infect. Dis. Soc. Am. 66, 1631 (2018).

15. Rhee, C., Strich, J. R., Klompas, M., Yealy, D. M. & Masur, H. Sep-1 has brought much needed attention to improving sepsis care… but now is the time to improve sep-1. Critical Care Medicine 48, 779–782 (2020).

16. Zhang, D., Micek, S. T. & Kollef, M. H. Time to appropriate antibiotic therapy is an independent determinant of postinfection icu and hospital lengths of stay in patients with sepsis. Critical care medicine 43, 2133–2140 (2015).

17. Shashikumar, S. P., Josef, C., Sharma, A. & Nemati, S. Deepaise–an end-to-end development and deployment of a recurrent neural survival model for early prediction of sepsis. arXiv preprint 1908.04759 (2019).

18. Tsoukalas, A., Albertson, T. & Tagkopoulos, I. From data to optimal decision making: a data-driven, probabilistic machine learning approach to decision support for patients with sepsis. JMIR medical informatics 3, e3445 (2015).

19. Raghu, A. et al. Deep reinforcement learning for sepsis treatment. arXiv preprint 1711.09602 (2017).

20. Raghu, A., Komorowski, M. & Singh, S. Model-based reinforcement learning for sepsis treatment. arXiv preprint 1811.09602 (2018).

21. Komorowski, M., Celi, L. A., Badawi, O., Gordon, A. C. & Faisal, A. A. The artificial intelligence clinician learns optimal treatment strategies for sepsis in intensive care. Nat. medicine 24, 1716–1720 (2018).

22. Utomo, C. P., Li, X. & Chen, W. Treatment recommendation in critical care: A scalable and interpretable approach in partially observable health states. In International Conference On Information Systems (2018).

23. Gal, Y. & Ghahramani, Z. Dropout as a bayesian approximation: Representing model uncertainty in deep learning. ICML’16 (JMLR.org, 2016).

24. Johnson, A. E. et al. Mimic-iii, a freely accessible critical care database. Sci. data 3, 1–9 (2016).

25. Thoral, P. J. et al. Sharing icu patient data responsibly under the society of critical care medicine/european society of intensive care medicine joint data science collaboration: The amsterdam university medical centers database (amsterdamumcdb) example. Critical care medicine 49, e563 (2021).

26. Hochreiter, S. & Schmidhuber, J. Long short-term memory. Neural computation 9, 1735–1780 (1997).

27. Rosenbaum, P. R. & Rubin, D. B. The central role of the propensity score in observational studies for causal effects. Biometrika 70, 41–55 (1983).

28. Singer, M. et al. The third international consensus definitions for sepsis and septic shock (sepsis-3). Jama 315, 801–810 (2016).

29. Vincent, J.-L. et al. The sofa (sepsis-related organ failure assessment) score to describe organ dysfunction/failure (1996).

30. Vincent, J.-L. et al. Use of the sofa score to assess the incidence of organ dysfunction/failure in intensive care units: results of a multicenter, prospective study. Critical care medicine 26, 1793–1800 (1998).

31. Ferreira, F. L., Bota, D. P., Bross, A., Mélot, C. & Vincent, J.-L. Serial evaluation of the sofa score to predict outcome in critically ill patients. Jama 286, 1754–1758 (2001).

32. Wager, S. & Athey, S. Estimation and inference of heterogeneous treatment effects using random forests. J. Am. Stat. Assoc. 113, 1228–1242 (2018).

33. Shalit, U., Johansson, F. D. & Sontag, D. Estimating individual treatment effect: generalization bounds and algorithms. In International Conference on Machine Learning, 3076–3085 (PMLR, 2017).

34. Seber, G. A. & Lee, A. J. Linear regression analysis, vol. 329 (John Wiley & Sons, 2012).

35. Liaw, A., Wiener, M. et al. Classification and regression by randomforest. R news 2, 18–22 (2002).

36. Wang, L., Zhang, Z. & Design, C. X. R. C. Theory and applications. Support. Vector Mach. Springer-Verlag, Berlin Heidelber (2005).

37. Hill, J. L. nBayesian nonparametric modeling for causal inference. J. Comput. Graph. Stat. 20, 217–240 (2011).

38. Yoon, J., Jordon, J. & van der Schaar, M. Ganite: Estimation of individualized treatment effects using generative adversarial nets. In International Conference on Learning Representations (2018).

39. Shi, C., Blei, D. & Veitch, V. Adapting neural networks for the estimation of treatment effects. In NeurIPS’19, 2503–2513 (2019).

40. Lim, B. Forecasting treatment responses over time using recurrent marginal structural networks. In NeurIPS’18, 7483–7493 (2018).

41. Bica, I., Alaa, A. M., Jordon, J. & van der Schaar, M. Estimating counterfactual treatment outcomes over time through adversarially balanced representations. arXiv preprint 2002.04083 (2020).

42. Dupuis, C. & Timsit, J.-F. Antibiotics in the first hour: is there new evidence? Expert. Rev. Anti-infective Ther. 19, 45–54 (2021).

43. Im, Y. et al. Time-to-antibiotics and clinical outcomes in patients with sepsis and septic shock: a prospective nationwide multicenter cohort study. Critical Care 26, 1–10 (2022).

44. Alam, N. et al. Prehospital antibiotics in the ambulance for sepsis: a multicentre, open label, randomised trial. The Lancet Respir. Medicine 6, 40–50 (2018).

45. Rhee, C. et al. Prevalence of antibiotic-resistant pathogens in culture-proven sepsis and outcomes associated with inadequate and broad-spectrum empiric antibiotic use. JAMA Netw. Open 3, e202899–e202899 (2020).

46. Singer, M. Antibiotics for sepsis: does each hour really count, or is it incestuous amplification? (2017).

47. Strich, J. R., Heil, E. L. & Masur, H. Considerations for empiric antimicrobial therapy in sepsis and septic shock in an era of antimicrobial resistance. The J. Infect. Dis. 222, S119–S131 (2020).

48. Severe sepsis and septic shock antibiotic guide - stanford medicine.

49. Rubin, D. B. Causal inference using potential outcomes: Design, modeling, decisions. J. Am. Stat. Assoc. 100, 322–331 (2005).

50. Robins, J. M. & Hernán, M. A. Estimation of the causal effects of time-varying exposures. Longitud. data analysis 553, 599 (2009).

51. Robins, J. M., Hernan, M. A. & Brumback, B. Marginal structural models and causal inference in epidemiology (2000).

52. Hernan, M. A. & Robins, J. M. Causal inference (2010).

53. Schwab, P., Linhardt, L. & Karlen, W. Perfect match: A simple method for learning representations for counterfactual inference with neural networks. arXiv preprint 1810.00656 (2018).

54. Jesson, A., Mindermann, S., Shalit, U. & Gal, Y. Identifying causal-effect inference failure with uncertainty-aware models. Adv. Neural Inf. Process. Syst. 33 (2020).

55. Bangaru, S. P., Suhas, J. & Ravindran, B. Exploration for multi-task reinforcement learning with deep generative models. arXiv preprint 1611.09894 (2016).

56. Xiao, T. & Kesineni, G. Generative adversarial networks for model based reinforcement learning with tree search. Univ. California, Berkeley, Tech. report (2016).

57. Andersen, P.-A., Goodwin, M. & Granmo, O.-C. The dreaming variational autoencoder for reinforcement learning environments. In International Conference on Innovative Techniques and Applications of Artificial Intelligence, 143–155 (Springer, 2018).

58. Bica, I., Alaa, A. M. & van der Schaar, M. Time series deconfounder: Estimating treatment effects over time in the presence of hidden confounders. arXiv preprint 1902.00450 (2019).

59. Li, R. et al. G-net: a recurrent network approach to g-computation for counterfactual prediction under a dynamic treatment regime. In Machine Learning for Health, 282–299 (PMLR, 2021).

60. Austin, P. C., Grootendorst, P. & Anderson, G. M. A comparison of the ability of different propensity score models to balance measured variables between treated and untreated subjects: a monte carlo study. Stat. medicine 26, 734–753 (2007).

61. Imbens, G. W. & Rubin, D. B. Causal inference in statistics, social, and biomedical sciences (Cambridge University Press, 2015).

62. Chandra, J. et al. A novel vascular leak index identifies sepsis patients with a higher risk for in-hospital death and fluid accumulation. Critical Care 26, 1–10 (2022).

