## Supplementary Material for "Estimating Treatment Effects for Time-to-Treatment Antibiotic Stewardship in Sepsis"

### Additional related work

Recently, many work of on-policy learning with reinforcement learning are proposed to first represent the actual environment by using a generative model and then learn optimal policies based on the generative environment. Bangaru et al.,[55] introduce an adaptive exploration signal as pseudo-reward from a deep generative model in order to deduce the Markov Decision Process (MDP). Xiao et al.,[56] propose to apply generative adversarial networks (GAN) to learn dynamics of the environment for model-based reinforcement learning. Andersen et al.,[57] propose to create a generative environment using variational autoencoder (VAE) and learn optimal policies based on the generated samples.

### Additional details on experimental setups

#### *Real-world data*

**Table 1.** The list of antibiotics in MIMIC-III Dataset. There are 18 kinds of antibiotics in total.

| Category | Name |
|---|---|
| Antibiotic | Cefazolin, Cefepime, Ceftazidime, Ciprofloxacin, Clindamycin, Erythromycin, Gentamicin, Levofloxacin, Metronidazole, Moxifloxacin, Piperacillin, Rifampin, Tobramycin, Vancomycin, Amikacin, Ampicillin, Azithromycin, Aztreonam |

**Table 2.** The definition of SOFA score and its components across six organ systems. Each SOFA component score ranges from 0 (normal) to 4 (most abnormal). The total SOFA score ranges from 0 (normal) to 24 (most abnormal).

| SOFA score | 1 | 2 | 3 | 4 |
|---|---|---|---|---|
| Respiration $PaO_2/FiO_2$, mmHg | $< 400$ | $< 300$ | $< 200$ | $< 100$ |
| Coagulation Platelets $\times 10^3$ /mm$^3$ | $< 150$ | $< 100$ | $< 50$ | $< 20$ |
| Liver Bilirubin, mg/dl ($\mu$mol/l) | 1.2 - 1.9 (20 - 32) | 2.0 - 5.9 (33 - 101) | 6.0 - 11.9 (102 - 204) | $> 12.0$ ($> 204$) |
| Cardiovascular Hypotension | MAP $< 70$ mmHg | Dopamine $\leq 5$ or dobutamine (any dose) | Dopamine $> 5$ or epinephrine $\leq 0.1$ or norepinephrine $\leq 0.1$ | Dopamine $> 15$ or epinephrine $> 0.1$ or norepinephrine $> 0.1$ |
| Central nervous system (CNS) Glasgow Coma Score (GCS) | 13 - 14 | 10 - 12 | 6 - 9 | $<6$ |
| Renal Creatinine, mg/dl ($\mu$mol/l) or urine output | 1.2 - 1.9 (110 - 170) | 2.0 - 3.4 (171 - 299) | 3.5-4.9 (300 - 440) or $< 500$ ml/day | $> 5.0$ ($> 440$) or $<200$ ml/day |

**Table 3.** The list of variables in MIMIC-III and AdmsterdamDB. There are 22 temporal covariates and 4 demographics and static variables. PT: Prothrombin Time; BUN: Blood Urea Nitrogen; WBC: White Blood Cells count;

| Category | | MIMIC-III | | AmsterdamDB | |
|---|---|---|---|---|---|
| | | Mean | *Std.* | Mean | *Std.* |
| Demographics | Age | 65.55 | 16.44 | 61.30 | 17.90 |
| | Gender | 43% Female | - | 42% Female | - |
| | Weight | 81.67 | 25.50 | 79.83 | 13.61 |
| | Height | 169.30 | 11.17 | 175.15 | 8.44 |
| Lab test | Anion gap | 13.35 | 3.80 | 8.70 | 4.62 |
| | Bicarbonate | 25.65 | 5.27 | 25.63 | 6.35 |
| | Bilirubin | 3.36 | 6.41 | 3.15 | 6.85 |
| | Creatinine | 1.50 | 1.46 | 1.28 | 1.03 |
| | Chloride | 104.00 | 6.60 | 108.60 | 46.31 |
| | Glucose | 134.00 | 66.83 | 133.9 | 45.74 |
| | Hematocrit | 29.96 | 5.13 | 38.98 | 1.67 |
| | Hemoglobin | 10.09 | 1.79 | 12.57 | 1.64 |
| | Lactate | 2.44 | 2.14 | 2.40 | 2.95 |
| | Platelet | 235.05 | 155.28 | 220.82 | 171.65 |
| | Potassium | 4.08 | 0.63 | 5.58 | 602.56 |
| | PT | 17.76 | 8.95 | 1.59 | 10.12 |
| | Sodium | 138.84 | 5.32 | 140.88 | 43.45 |
| | BUN | 29.85 | 23.54 | 14.15 | 9.80 |
| | WBC | 11.23 | 7.64 | 14.56 | 11.80 |
| Vital signs | Heart Rate | 87.81 | 18.30 | 92.70 | 23.65 |
| | SysBP | 120.92 | 23.28 | 126.05 | 139.59 |
| | DiasBP | 61.41 | 14.55 | 60.77 | 31.11 |
| | Meanbp | 78.70 | 16.88 | 82.12 | 47.34 |
| | Respratory | 20.48 | 5.90 | 21.99 | 7.71 |
| | Temperature | 36.96 | 0.85 | 36.73 | 21.14 |
| | SpO2 | 97.00 | 3.27 | 96.09 | 7.43 |

```
Seq2Seq(
  [illegible]: Encoder(
    ([illegible]ing): Linear(in_features=22, out_features=32, bias=True)
    (embedding_static): Linear(in_features=4, out_features=32, bias=True)
    (rnn): LSTM(33, 64, num_layers=2, batch_first=True)
    (fc_out): Linear(in_features=97, out_features=1, bias=True)
    (dropout): Dropout(p=0.3, inplace=False)
    (attention_encoder): Sequential(
      (0): Linear(in_features=64, out_features=1, bias=True)
      (1): Tanh()
    )
    (ps_out): Sequential(
      (0): Linear(in_features=96, out_features=64, bias=True)
      (1): ReLU()
      (2): Linear(in_features=64, out_features=1, bias=False)
    )
  )
  (decoder): AttentionDecoder(
    (embedding): Linear(in_features=1, out_features=32, bias=True)
    (embedding_static): Linear(in_features=4, out_features=32, bias=True)
    (attn_f): Attn(
      (attn): Linear(in_features=64, out_features=64, bias=True)
    )
    (rnn): LSTM(65, 64, num_layers=2, batch_first=True, dropout=0.3)
    (fc_out_1): Sequential(
      (0): Linear(in_features=128, out_features=64, bias=True)
      (1): ReLU()
      (2): Linear(in_features=64, out_features=1, bias=True)
    )
    (fc_out_0): Sequential(
      (0): Linear(in_features=128, out_features=64, bias=True)
      (1): ReLU()
      (2): Linear(in_features=64, out_features=1, bias=True)
    )
    (ps_out): Sequential(
      (0): Linear(in_features=128, out_features=64, bias=True)
      (1): ReLU()
      (2): Linear(in_features=64, out_features=1, bias=True)
    )
    (dropout): Dropout(p=0.3, inplace=False)
  )
)
```

Scroll to End

**Figure 1.** Model configuration.

**Table 4.** Hyperparameter search range and optimal hyperparameters for **T4**.

| | Hyperparameter range | Optimal hyperparameters |
|---|---|---|
| Augmentation ratio | 0, 0.1, 0.2, 0.3, 0.4, 0.5, 0.6, 0.7, 0.8, 0.9 1.0 | 0.4 |
| Learning rate | 5e-3, 1e-3, 5e-4, 1e-4, 5e-5, 1e-5 | 5e-5 |
| Embedding size | 16, 32, 64, 128 | 32 |
| Hidden layer size | 32, 64, 128, 256 | 128 |
| Dropout rate | 0.1, 0.2, 0.3, 0.4, 0.5 | 0.3 |
| Batch size | 8, 16, 32, 64 | 32 |

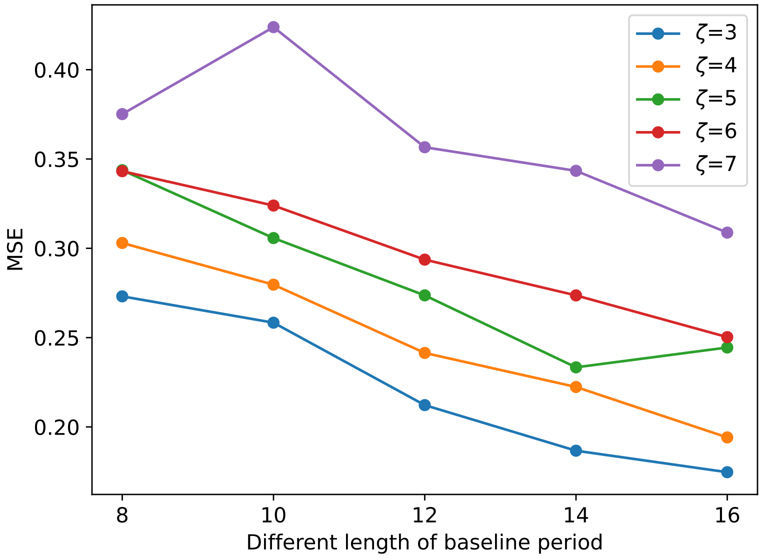


**Figure 2.** The influence of different length of baseline period to factual prediction performance among different length of follow-up period on MIMIC-III dataset.

#### *Synthetic Data*

To illustrate the model performance on ITE estimation and treatment recommendation, we design experiments on a synthetic dataset. We first simulate temporal covariates $x_t$ as a weighted sum of historical covariates $\overline{x}_{t-1}$ and treatment assignments $\overline{a}_{t-1}$ as follows,

$$x_t|\overline{x}_{t-1},\overline{a}_{t-1} \sim \frac{1}{\sum_{j=1}^{t-1} w_j} \sum_{j=1}^{t-1} w_j(x_j + \lambda a_j) \tag{21}$$

where the weight $w_j = w_{j-1} * 2$ forms a geometric sequence with ratio 2, $\lambda \sim \mathcal{N}(0, 0.1)$ controls the influence of historical treatment assignments. The initial covariates are simulated by a multi-variable Gaussian distribution as $x_1 \sim \mathcal{N}(0^{k_t}, 0.1 \cdot (\Sigma \cdot \Sigma^\top))$, where $\Sigma \sim \mathscr{U}((-1,1)^{k_t \times k_t})$ is simulated by a uniform distribution and $k_t$ is the dimension of the temporal covariates. We also simulate static covariates $c$ with the same distribution. We then simulate the treatment assignment $a_t$ at each timestamp as,

$$a_t|x_t, c \sim \texttt{Bernoulli}(\sigma(s^\top [x_t, c] + m)) \tag{22}$$

where $s \sim \mathcal{N}(0^k, 0.1 \cdot I)$, $k$ is the dimension of all covariates, $I$ is the identity matrix, and $m \sim \mathcal{N}(0, 0.1)$. $\sigma(\cdot)$ is the sigmoid function and $[\cdot]$ concatenates two vectors as a whole. The outcome is simulated as a function of temporal covariates, static covariates and treatments as follows,

$$y_t|x_t, a_t \sim q^\top [x_t, c, \beta a_t] + \varepsilon \tag{23}$$

where $q \sim \mathcal{N}(0^{k+1}, 0.05 \cdot (\Sigma \cdot \Sigma^\top))$, $\Sigma \sim \mathscr{U}((-1,1)^{(k+1)\times(k+1)})$, $\beta \sim \mathcal{N}(0, 0.5)$ controls the influence of treatments, and $\varepsilon \sim \mathcal{N}(0, 0.1)$.

In this paper, we simulate 5000 patients with 50 timestamps, $k_t = 20$ temporal covariates and 5 static covariates. We use the first 40 timestamps as baseline period and the remaining as follow-up period. Note that the timestamp here corresponds to the hour in the physical time unit without any time interval aggregation as in real-world data. The synthetic dataset and codes for simulation are available at Github[5].

#### *Baseline Methods*

We conduct comparison experiments against the state-of-the-art methods of ITE estimation in the following categories: (1) **Classical methods:** Linear Regression (LR)[34], Random Forest (RF)[35] and support vector machine (SVM)[36] are included as the basic machine learning models for comparison. They directly regard the treatment assignment as an additional feature and predict potential outcomes based on the patient's covariates under different treatments; (2) **Forest-based methods:** Causal Forest (CF)[32] and Bayesian Additive Regression Trees (BART)[37] are two commonly used tree-based models for causal effect estimation. CF extends classical model RF that instead of minimizing prediction error, it split the data for maximizing the difference of treatment effects across splits. BART is a non-parametric Bayesian regression tree model that each tree is a learner constrained by a regularization prior; (3) **Representation learning-based methods:** Counterfactual Regression (CFR)[33] construct balanced representations in the hidden space via deep neural network. We compare two variants of CFR: one is equipped with Wasserstein (WASS) as distance metrics for distribution balance and the other is the vanilla version without any distribution balance (TARNET). GANITE[38] estimates the ITEs via a generative adversarial network by generating and discriminating counterfactual. Dragonnet[39] jointly optimize propensity prediction and potential outcome prediction for ITE estimation. (4) **Time-varying based methods: Recurrent Marginal structural Network (RMSM)[40]** adopts recurrent marginal structural network for predicting the patient's potential response to a series of treatments. Counterfactual Recurrent Network (CRN)[41] adopts adversarial training techniques to balance the historical confounding variables.

#### *Performance Measurement*

We use Precision in Estimation of Heterogeneous Effect (PEHE) to evaluate the model performance on ITE estimation. Specifically, PEHE computes the mean squared error (MSE) between the values of ground truth ITE $\delta^i_j$ and estimated ITE $\delta'^i_j$ as follows,

$$\text{PEHE} = \frac{1}{\zeta}\frac{1}{N}\sum_{j=1}^{\zeta}\sum_{i=1}^{N}(\delta^i_j - \delta'^i_j)^2 \tag{24}$$

[5] https://github.com/ruoqi-liu/T4

Besides the individual-level evaluation, we are also interested in the causal effect over the entire population. We use the error of Average Treatment Effect ($\varepsilon$ATE) to evaluate the model performance, which is computed as the mean absolute error (MAE) between the ground truth and estimated ATE as,

$$\varepsilon \text{ATE} = \frac{1}{\zeta} \sum_{j=1}^{\zeta} \left| \frac{1}{N} \sum_{i=1}^{N} \delta_j^i - \frac{1}{N} \sum_{i=1}^{N} \delta_j'^i \right| \tag{25}$$

Two metrics are all averaged on $\zeta$ follow-up timestamps and PEHE is regarded as the primary evaluation metric.

### Additional experimental results

#### *Real-world data*

**Table 5.** The average standard deviation of estimated treatment effect $\delta$ in the follow-up period.

| $\zeta$ | 1 | 2 | 3 | 4 | 5 | 6 |
|---|---|---|---|---|---|---|
| Standard deviation of $\delta$ | 0.03843 | 0.03950 | 0.03975 | 0.03996 | 0.03992 | 0.04011 |

**Table 6.** The percentage of patients who received treatments different from model's recommendation (median, %).

| | 30-day mortality | | | | | 60-day mortality | | | | |
|---|---|---|---|---|---|---|---|---|---|---|
| Follow-up period | 3 | 4 | 5 | 6 | 7 | 3 | 4 | 5 | 6 | 7 |
| MIMIC-III | 50.10 | 53.40 | 48.81 | 36.37 | 23.90 | 50.41 | 53.24 | 50.38 | 36.39 | 23.69 |
| AmsterdamUMCdb | 54.55 | 64.28 | 67.46 | 64.72 | 59.33 | 54.76 | 64.34 | 68.17 | 63.00 | 58.58 |

**Table 7.** Performance comparison of factual prediction of SOFA scores on MIMIC-III dataset. Here, we report the estimated mean squared error (MSE) of each method among five different lengths of follow-up period.

| | Method | $\zeta=3$ | $\zeta=4$ | $\zeta=5$ | $\zeta=6$ | $\zeta=7$ |
|---|---|---|---|---|---|---|
| Base model | LR | $0.739 \pm 0.094$ | $0.749 \pm 0.099$ | $0.768 \pm 0.122$ | $0.803 \pm 0.171$ | $0.841 \pm 0.233$ |
| | RF | $0.613 \pm 0.030$ | $0.624 \pm 0.029$ | $0.634 \pm 0.031$ | $0.638 \pm 0.032$ | $0.635 \pm 0.028$ |
| | SVM | $0.639 \pm 0.029$ | $0.646 \pm 0.030$ | $0.652 \pm 0.031$ | $0.656 \pm 0.030$ | $0.660 \pm 0.031$ |
| Representation learning based | CFR WASS[33] | $0.646 \pm 0.017$ | $0.657 \pm 0.019$ | $0.656 \pm 0.015$ | $0.665 \pm 0.018$ | $0.674 \pm 0.015$ |
| | TARNET[33] | $0.663 \pm 0.012$ | $0.682 \pm 0.022$ | $0.697 \pm 0.014$ | $0.687 \pm 0.017$ | $0.688 \pm 0.024$ |
| | GANITE[38] | $0.877 \pm 0.016$ | $0.884 \pm 0.019$ | $0.892 \pm 0.013$ | $0.896 \pm 0.014$ | $0.904 \pm 0.013$ |
| | Dragonnet[39] | $0.642 \pm 0.012$ | $0.640 \pm 0.009$ | $0.650 \pm 0.015$ | $0.665 \pm 0.022$ | $0.666 \pm 0.017$ |
| Forest based | Causal Forest[32] | $0.657 \pm 0.022$ | $0.666 \pm 0.022$ | $0.676 \pm 0.020$ | $0.682 \pm 0.021$ | $0.681 \pm 0.020$ |
| | BART[37] | $0.579 \pm 0.028$ | $0.594 \pm 0.024$ | $0.612 \pm 0.028$ | $0.617 \pm 0.022$ | $0.619 \pm 0.024$ |
| Time-varying based | RMSN[40] | $0.247 \pm 0.020$ | $0.233 \pm 0.007$ | $0.260 \pm 0.005$ | $0.321 \pm 0.017$ | $0.378 \pm 0.017$ |
| | CRN[58] | $0.225 \pm 0.011$ | $0.235 \pm 0.008$ | $0.263 \pm 0.007$ | $0.319 \pm 0.018$ | $0.391 \pm 0.015$ |
| | G-Net[59] | $0.221 \pm 0.010$ | $0.230 \pm 0.006$ | $0.261 \pm 0.009$ | $0.313 \pm 0.008$ | $0.372 \pm 0.012$ |
| Ours | **T4** | $\mathbf{0.173 \pm 0.013}$ | $\mathbf{0.212 \pm 0.011}$ | $\mathbf{0.248 \pm 0.007}$ | $\mathbf{0.275 \pm 0.013}$ | $\mathbf{0.335 \pm 0.019}$ |

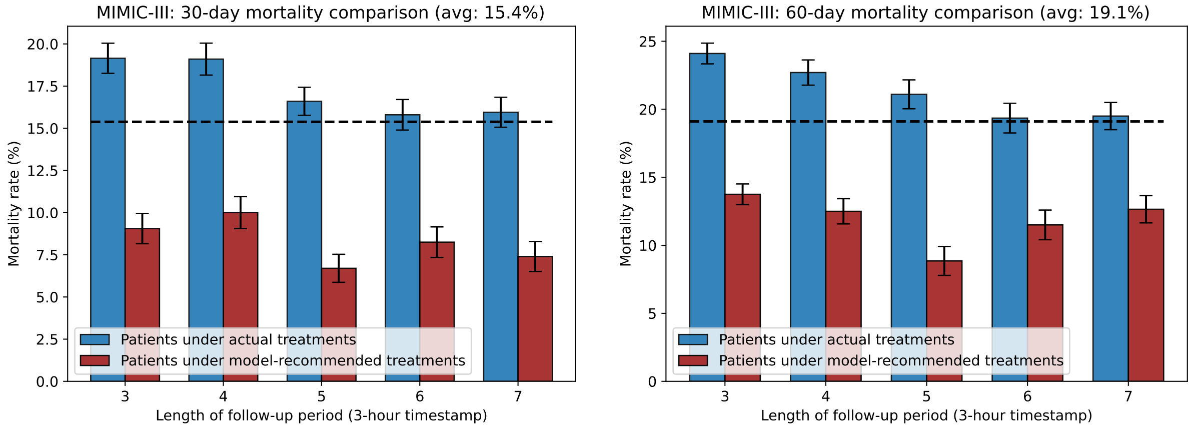


(a) (b)

**Figure 3.** Mortality rate comparison of suspected septic patients obtained from MIMIC-III dataset. Within the bar-chart, the error bars denote 95% confidence interval with $n$=30 bootstrap samples. Blue and red bars denote patients under actual treatments and patients under model-recommended treatments, respectively.

The total mortality rate of two groups of patients is plotted using black dashed line, which serves as the baseline.

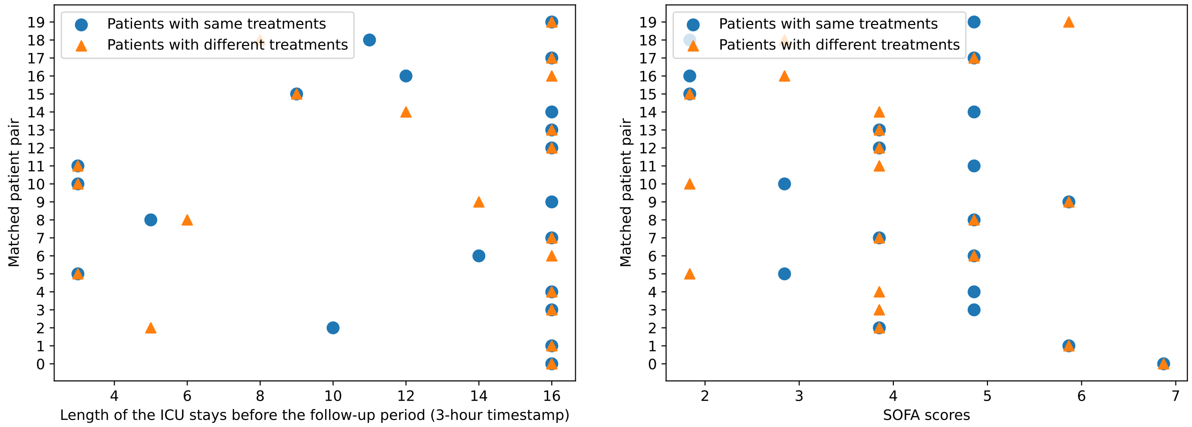


(a) (b)

**Figure 4.** Matching evaluation of 20 randomly selected patients with different treatment timing as recommendation (yellow triangles) and their matched pairs with same treatment timing (blue circles). Fig.4(a) shows the length of baseline period of matched pairs. Fig. 4(b) shows the SOFA scores of matched pairs.

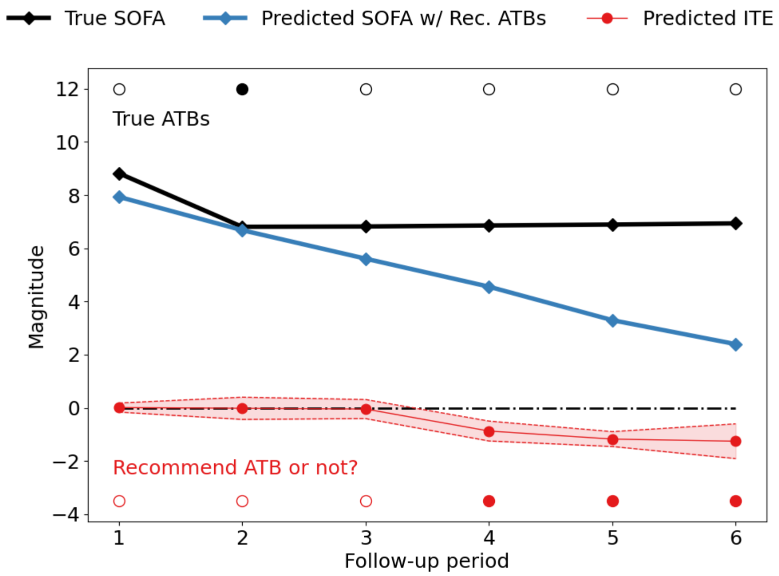


**Figure 5.** An example to illustrate the treatment recommendation process. The predicted values of ITEs (red line with shadowed area denoting the uncertainty estimates) for antibiotic (ATB) recommendation. An ATB will be recommended to the patient if the upper bound of predicted ITE is lower than zero and will not be recommended if the lower bound of predicted ITE is higher than zero, where zero is the threshold for determining whether to recommend ATBs. The patient takes ATB in early stage of the follow-up period, while the model recommends to take ATB later. The predicted SOFA score under recommended ATBs (blue) is much lower than the true SOFA score (black line).

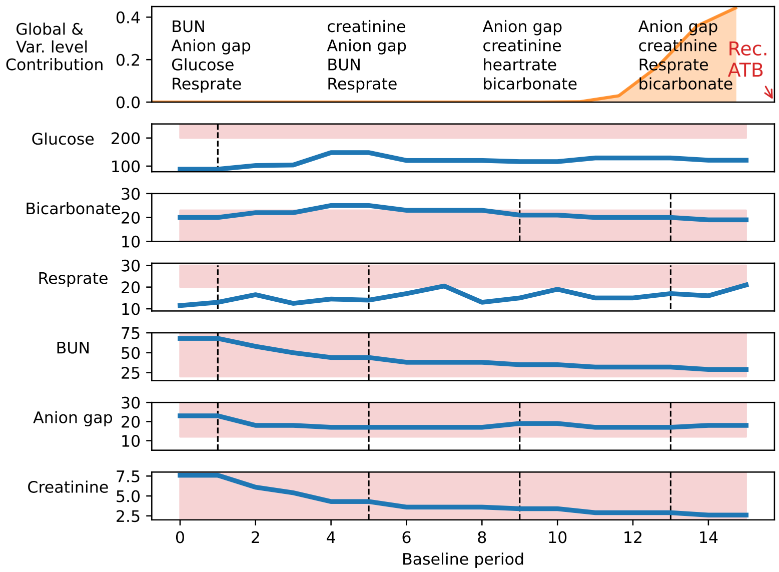


**Figure 6.** An example to illustrate model interpretability on treatment recommendation. The most important global baseline timestamps (orange area) and variables contributing to the treatment recommendation are denoted at the top subplot. An ATB is recommended at the end of the baseline period. **T4** provides transparent antibiotics recommendation based on the time-varying variables in the baseline period.

#### ***Synthetic data***

The performance of the proposed **T4** model and existing baseline models on different lengths of follow-up period is shown in Supplementary Table 8. We vary $\zeta = 3$ to 7 and observe that **T4** demonstrates better performance compared to the baselines in terms of PEHE and $\varepsilon$ATE. We first examine the performance of base machine learning models including LR, RF and SVM. They directly regard the treatment assignment as an additional feature without considering the influence of confounders. Thus, the overall performance is lower than the other two categories which adjusts the influence of confounders and selection bias. Among the three base models, SVM performs better than others for different lengths of the follow-up period.

Representation learning based approaches achieve better performance than the base models. They take advantage of deep neural network to learn the representations of confounders. GANITE, which adopts the Generative Adversarial Nets (GANs) for estimating ITEs by modeling the counterfactual distributions, performs better than other representation learning models. TARNET and CFR WASS share the same fundamental framework excepts that CFR WASS is equipped with Wasserstein (WASS) as distance measurement for balancing the distribution of different treatment groups in the hidden space. Thus, the performance of CFR WASS is better than TARNET. Dragonnet achieves comparative performance with TARNET. The performance of forest based models is relatively better than representation learning based models and BART achieves better scores than the CF. Time-varying based methods (RMSN and CRN) are designed to estimate ITE with time-varying data. They generally outperform other baselines which are mainly designed for static data.

Our proposed **T4** achieves outstanding performance than all baselines. The model fully considers the temporality of data by modeling the time-varying confounders via a recurrent neural network, and adjusting the influence of confounders by constructing *pseudo* balanced training batches via balancing matching operation. To evaluate the effectiveness of balancing matching, we design two ablations of the original model. **1) T4-w/o BM:** we remove the entire balancing matching component from the model; **2) T4-w/ Rand.:** we augment the training batches with a portion of randomly selected samples. From the Supplementary Table 8, we observe the performance of **T4**-w/o BM is lower than the complete **T4** model, which demonstrates that balancing matching improves the model performance by constructing balanced batches and adjusting confounders. We also find that the performance of **T4**-w/ Rand. is better than **T4**-w/o BM but far from **T4**. The results show that random mini-batch augmentation only yields limited improvement. Our proposed balancing matching is specifically designed to adjust the influence of confounding via matching the samples based on the computed balancing scores, and therefore fundamentally facilitates the ITE estimation. Thus, compared to **T4**-w/o BM and **T4**-w/ Rand., **T4** achieves the best performance for ITE estimation.

We also consider an enhancing MC-dropout with variational dropout for recurrent networks[23] as a replacement for the original dropout layer (i.e., dropout is applied on the embedding layer). The performance of our model with variational dropout (**T4** (Variat. MC)) is comparable to the model with original dropout layer.

#### ***Influence of uncertainty quantification to treatment recommendation***

We use the estimated uncertainty to quantify the confidence of estimated ITEs and provided treatment recommendation. In Supplementary Fig. 7, we demonstrate that our model equipped with uncertainty estimates $\eta(\delta')$ achieves lower estimation error when excluding fewer individuals (withholding recommendation) with uncertain estimation compared to other uncertainty quantification approaches. Here, the percentage of recommendation withheld in the figure denotes the percentage of excluded individuals with low confidence during the evaluation. The approach of propensity[60] computes and rank the propensity scores (Eq. (12)) for each individual. Then it excludes the individuals with computed propensity scores either close to 0 or 1. These individuals are likely to violate the overlap assumption[61] in causal inference and lead to inaccurate ITE estimation. The random approach randomly excludes the individuals and serves as the baseline.

We vary the percentage of recommendation withheld and show the PEHE and $\varepsilon$ATE on the remaining data samples in the figure. We observe that estimated PEHE and $\varepsilon$ATE for three approaches all decline with the increasing percentage of recommendation withheld, because samples with high uncertainty and inaccurate ITE estimation are excluded sequentially. The uncertainty quantification method $\eta(\delta')$ we adopt in our model achieves the best performance among others in terms of fast decreasing estimation error when the percentage of recommendation withheld increases. The results demonstrate that uncertainty quantification helps to treatment recommendation by explicitly examining the confidence associated with ITE estimation and treatment recommendation.

**Table 8.** Performance comparison of ITE estimation on synthetic datasets. Here, we report the estimated PEHE and $\varepsilon$ATE of each method among five different lengths of follow-up period.

| Method | | $\zeta = 3$ | | $\zeta = 4$ | | $\zeta = 5$ | | $\zeta = 6$ | | $\zeta = 7$ | |
|---|---|---|---|---|---|---|---|---|---|---|---|
| | | PEHE | $\varepsilon$ATE | PEHE | $\varepsilon$ATE | PEHE | $\varepsilon$ATE | PEHE | $\varepsilon$ATE | PEHE | $\varepsilon$ATE |
| Base model | LR | 3.958 ± 0.072 | 1.500 ± 0.026 | 3.987 ± 0.072 | 1.505 ± 0.025 | 4.017 ± 0.07 | 1.511 ± 0.024 | 4.048 ± 0.064 | 1.517 ± 0.023 | 4.086 ± 0.059 | 1.524 ± 0.023 |
| | RF | 3.989 ± 0.077 | 1.506 ± 0.027 | 4.021 ± 0.077 | 1.512 ± 0.026 | 4.052 ± 0.076 | 1.518 ± 0.026 | 4.084 ± 0.069 | 1.523 ± 0.025 | 4.124 ± 0.062 | 1.53 ± 0.024 |
| | SVM | 3.947 ± 0.075 | 1.498 ± 0.027 | 3.979 ± 0.074 | 1.504 ± 0.026 | 4.007 ± 0.074 | 1.509 ± 0.026 | 4.038 ± 0.070 | 1.515 ± 0.025 | 4.078 ± 0.064 | 1.522 ± 0.025 |
| Representation learning based | CFR WASS[33] | 3.794 ± 0.199 | 1.447 ± 0.032 | 3.823 ± 0.188 | 1.451 ± 0.030 | 3.851 ± 0.175 | 1.455 ± 0.029 | 3.884 ± 0.178 | 1.459 ± 0.029 | 3.914 ± 0.178 | 1.464 ± 0.029 |
| | TARNET[33] | 3.848 ± 0.099 | 1.469 ± 0.020 | 3.898 ± 0.108 | 1.478 ± 0.021 | 3.937 ± 0.103 | 1.484 ± 0.021 | 3.967 ± 0.102 | 1.489 ± 0.021 | 3.998 ± 0.116 | 1.494 ± 0.023 |
| | GANITE[38] | 3.786 ± 0.212 | 1.460 ± 0.035 | 3.808 ± 0.195 | 1.463 ± 0.033 | 3.842 ± 0.195 | 1.471 ± 0.034 | 3.869 ± 0.208 | 1.473 ± 0.035 | 3.911 ± 0.204 | 1.480 ± 0.035 |
| | Dragonnet[39] | 3.830 ± 0.084 | 1.466 ± 0.018 | 3.872 ± 0.096 | 3.872 ± 0.096 | 3.922 ± 0.091 | 1.480 ± 0.019 | 3.945 ± 0.098 | 1.484 ± 0.020 | 3.980 ± 0.112 | 1.491 ± 0.022 |
| Forest based | Causal Forest[32] | 3.755 ± 0.201 | 1.441 ± 0.033 | 3.785 ± 0.192 | 1.446 ± 0.031 | 3.810 ± 0.180 | 1.449 ± 0.027 | 3.843 ± 0.182 | 1.454 ± 0.022 | 3.875 ± 0.182 | 1.459 ± 0.030 |
| | BART[37] | 3.750 ± 0.205 | 1.44 ± 0.034 | 3.782 ± 0.193 | 1.446 ± 0.031 | 3.806 ± 0.176 | 1.449 ± 0.030 | 3.841 ± 0.180 | 1.454 ± 0.030 | 3.871 ± 0.180 | 1.459 ± 0.029 |
| Time-varying based | RMSN[40] | 2.480 ± 0.022 | 1.195 ± 0.011 | 2.510 ± 0.017 | 1.201 ± 0.009 | 2.599 ± 0.032 | 1.218 ± 0.012 | 2.569 ± 0.027 | 1.211 ± 0.012 | 2.661 ± 0.027 | 1.230 ± 0.018 |
| | CRN[58] | 2.470 ± 0.030 | 1.187 ± 0.010 | 2.539 ± 0.024 | 1.202 ± 0.011 | 2.627 ± 0.028 | 1.219 ± 0.011 | 2.631 ± 0.031 | 1.219 ± 0.014 | 2.675 ± 0.029 | 1.235 ± 0.019 |
| | G-Net[59] | 2.476 ± 0.009 | 1.193 ± 0.014 | 2.507 ± 0.011 | 1.200 ± 0.009 | 2.591 ± 0.013 | 1.213 ± 0.007 | 2.558 ± 0.013 | 1.213 ± 0.011 | 2.651 ± 0.015 | 1.225 ± 0.012 |
| Ours | **T4**-w/o BM | 2.439 ± 0.012 | 1.175 ± 0.008 | 2.495 ± 0.019 | 1.188 ± 0.010 | 2.516 ± 0.017 | 1.192 ± 0.007 | 2.555 ± 0.021 | 1.200 ± 0.009 | 2.582 ± 0.024 | 1.207 ± 0.014 |
| | **T4**-w/ Rand. | 2.406 ± 0.016 | 1.165 ± 0.011 | 2.474 ± 0.015 | 1.183 ± 0.011 | 2.491 ± 0.013 | 1.186 ± 0.012 | 2.510 ± 0.019 | 1.189 ± 0.010 | 2.548 ± 0.020 | 1.197 ± 0.012 |
| | **T4** (Variat. MC) | 2.372 ± 0.014 | 1.160 ± 0.012 | 2.416 ± 0.020 | 1.168 ± 0.011 | 2.442 ± 0.021 | 1.167 ± 0.009 | 2.466 ± 0.019 | 1.174 ± 0.013 | 2.557 ± 0.026 | 1.200 ± 0.015 |
| | **T4** | **2.362 ± 0.020** | **1.149 ± 0.012** | **2.384 ± 0.019** | **1.156 ± 0.011** | **2.430 ± 0.022** | **1.164 ± 0.014** | **2.473 ± 0.019** | **1.174 ± 0.007** | **2.476 ± 0.024** | **1.176 ± 0.010** |

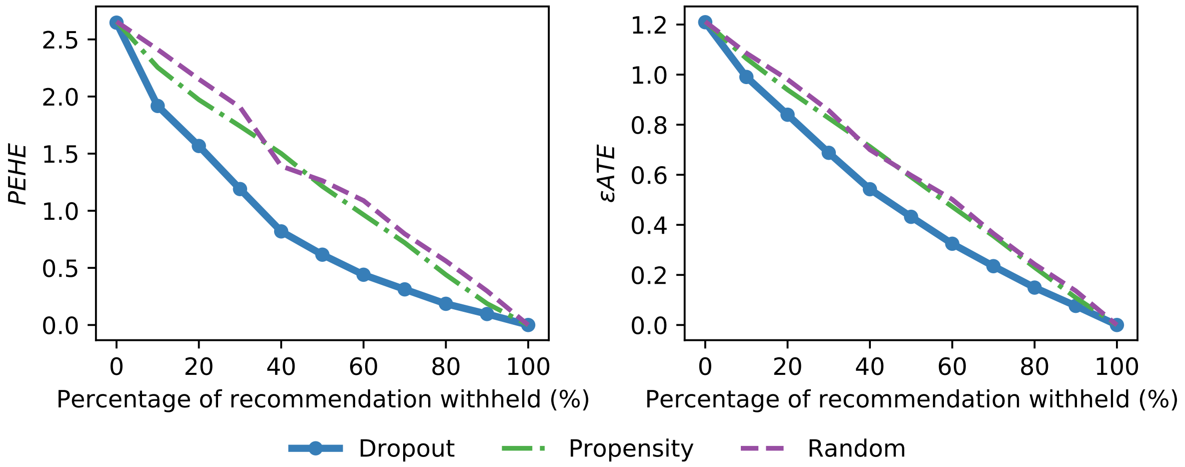


**Figure 7.** The estimated PEHE and $\varepsilon$ATE of different uncertainty quantification approaches when we vary the percentage of recommendation withheld. The evaluation metrics are reported on the remaining samples with provided recommendation.

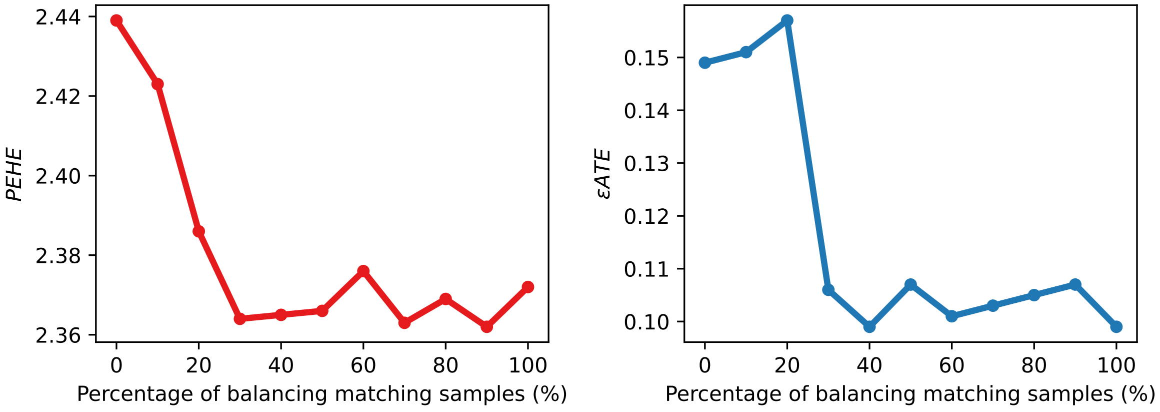


**Figure 8.** The performance change with regards to PEHE and $\varepsilon$ATE when the percentage of balancing matching samples in each training batch increases.
